# Clinical Knowledge Extraction via Sparse Embedding Regression (KESER) with Multi-Center Large Scale Electronic Health Record Data

**DOI:** 10.1101/2021.03.13.21253486

**Authors:** Chuan Hong, Everett Rush, Molei Liu, Doudou Zhou, Jiehuan Sun, Aaron Sonabend, Victor M. Castro, Petra Schubert, Vidul A. Panickan, Tianrun Cai, Lauren Costa, Zeling He, Nicholas Link, Ronald Hauser, J. Michael Gaziano, Shawn N. Murphy, George Ostrouchov, Yuk-Lam Ho, Edmon Begoli, Junwei Lu, Kelly Cho, Katherine P. Liao, Tianxi Cai, with the VA Million Veteran Program

## Abstract

**Objective:** The increasing availability of Electronic Health Record (EHR) systems has created enormous potential for translational research. Even with a working knowledge of EHR, it is difficult to know all the relevant codes related to a phenotype due to the large number of codes available. Traditional data mining approaches often require the use of patient-level data, which hinders the ability to share data across institutions to establish a cooperative and integrated knowledge network. In this project, we demonstrate that multi-center large-scale code embeddings can be used to efficiently identify relevant features related to a disease or condition of interest.

**Method:** We constructed large-scale code embeddings for a wide range of codified concepts, including diagnosis codes, medications, procedures, and laboratory tests from EHRs from two large medical centers. We developed knowledge extraction via sparse embedding regression (KESER) for feature selection and integrative network analysis based on the trained code embeddings. We evaluated the quality of the code embeddings and assessed the performance of KESER in feature selection for eight diseases. Besides, we developed an integrated clinical knowledge map combining embedding data from both institutions.

**Results:** The features selected by KESER were comprehensive compared to lists of codified data generated by domain experts. Additionally, features identified automatically via KESER used in the development of phenotype algorithms resulted in comparable performance to those built upon features selected manually or identified via existing feature selection methods with patient-level data. The knowledge map created using an integrative analysis identified disease-disease and disease-drug pairs more accurately compared to those identified using single institution data.

**Conclusion:** Analysis of code embeddings via KESER can effectively reveal clinical knowledge and infer relatedness among diseases, treatment, procedures, and laboratory measurement. This approach automates the grouping of clinical features facilitating studies of the condition. KESER bypasses the need for patient-level data in individual analyses providing a significant advance in enabling multi-center studies using EHR data.

## INTRODUCTION

The adoption of electronic health record (EHR) systems has simultaneously changed clinical practice and expanded the breadth of biomedical research. For clinical research studies, EHR data are used alone or integrated with other established data sources such as registries, genomic data from biobanks, and administrative databases^1–7^. EHR clinical data typically includes diagnostic billing codes, laboratory orders and results, procedure codes, and medication prescriptions. These comprehensive longitudinal data allow for studies to examine a broad range of hypotheses. However, this wealth of data also raises challenges in selecting and creating EHR features among thousands of options relevant to the study or condition of interest. Most current studies manually select individual EHR features and map specific EHR codes to represent each feature, requiring input from clinical and informatics experts. In addition to being susceptible to subjective bias, this manual, time-consuming process cannot be scaled for projects requiring multiple phenotypes.

Moreover, sharing algorithms across institutions often requires performing this manual process to identify institution-specific codes and coding patterns in collaborative or replication studies. One potential solution is to create large-scale clinical knowledge networks, providing information about the dependency structure across different EHR elements, thereby providing information about the relationship of conditions and codes at a particular institution as well as equivalent codes across institutions. These data would no longer be associated with individual patient data and could be readily shared, facilitating multi-center collaborations.

Creating a clinical knowledge network using EHR data requires two major advancements. First, a general approach is needed to integrate the different types of structured data efficiently, also referred to as codified data, available in EHR. Codified EHR data includes ICD (International Classification of Disease) codes^8,9^ for disease conditions, LOINC (Logical Observation Identifiers Names and Codes)^10^ for laboratory tests, CPT (Current Procedural Terminology)^11^ and CCS (Clinical Classifications Software)^12^ for procedures, as well as RxNorm^13^ and NDC (National Drug Code) for medications. Approaches for extracting knowledge from codified EHR data using machine learning algorithms have been proposed in recent years^14–16^. However, these algorithms focused on a specific task and required training with patient-level EHR data. Second, establishing a highly cooperative and shareable clinical knowledge network across institutions requires methods that can ensure data privacy. Existing approaches for data mining require patient-level EHR data, posing significant administrative challenges for data sharing across research groups and institutions.

To overcome these challenges, we propose to transform EHR data into embedding vectors^17^, thus uncoupling the data from the individual patient. The downstream machine learning tasks would use the embeddings vector as summary data rather than individual patient data. Our use of embedding in this study refers to projecting an EHR code into another representation space. In the past decade, embedding vectors have been successfully derived for clinical concepts with textual data and various sub-domains of codified EHR data^18–24^. These embeddings were primarily derived for specific applications and not for the creation of knowledge networks. In addition, most existing word embedding algorithms tuned the key hyper-parameters, e.g., the appropriate dimension of the embedding vectors, to optimize a specific downstream task. For example, the Code2Vec^19^ tuned the embedding dimension via clustering task, and the Med2Vec^21^ chose the dimension via future code prediction. However, this approach may limit the applicability of the learned embedding vectors to other downstream tasks. This study aims to develop a **<u>K</u>**nowledge **<u>E</u>**xtraction pipeline via **<u>S</u>**parse **<u>E</u>**mbedding **<u>R</u>**egression (KESER) with HER data from two large healthcare systems. We present methods to derive embedding vectors using multiple types of codified EHR data at scale. Here, we choose the hyperparameters to ensure the general quality of the embedding vectors and retain embedding vectors with higher dimensions to further enable users to fine-tune optimal dimensions for their specific tasks. We also investigate to what extent the dimensions affect the performance of different tasks. With embedding vectors from both institutions, we fit graphical models via sparse regression to construct knowledge networks that encode relatedness among features. We then demonstrate how these knowledge networks can select potential features in the development of an algorithm to identify patients with specific phenotypes using EHR data. Furthermore, we demonstrate that the knowledge network trained via integrative analysis of embedding data from both institutions outperforms those trained with a single institution’s data.

## RESULTS

The KESER procedure includes four key steps outlined in Figure 1: (i) data pre-processing; (ii) creating embedding vectors via representation learning using co-occurrence data and pointwise mutual information; (iii) feature selection at a single site via sparse regression; (iv) building a knowledge network across multiple sites via an integrative sparse regression and node-wise graphical model.

**Figure 1.**
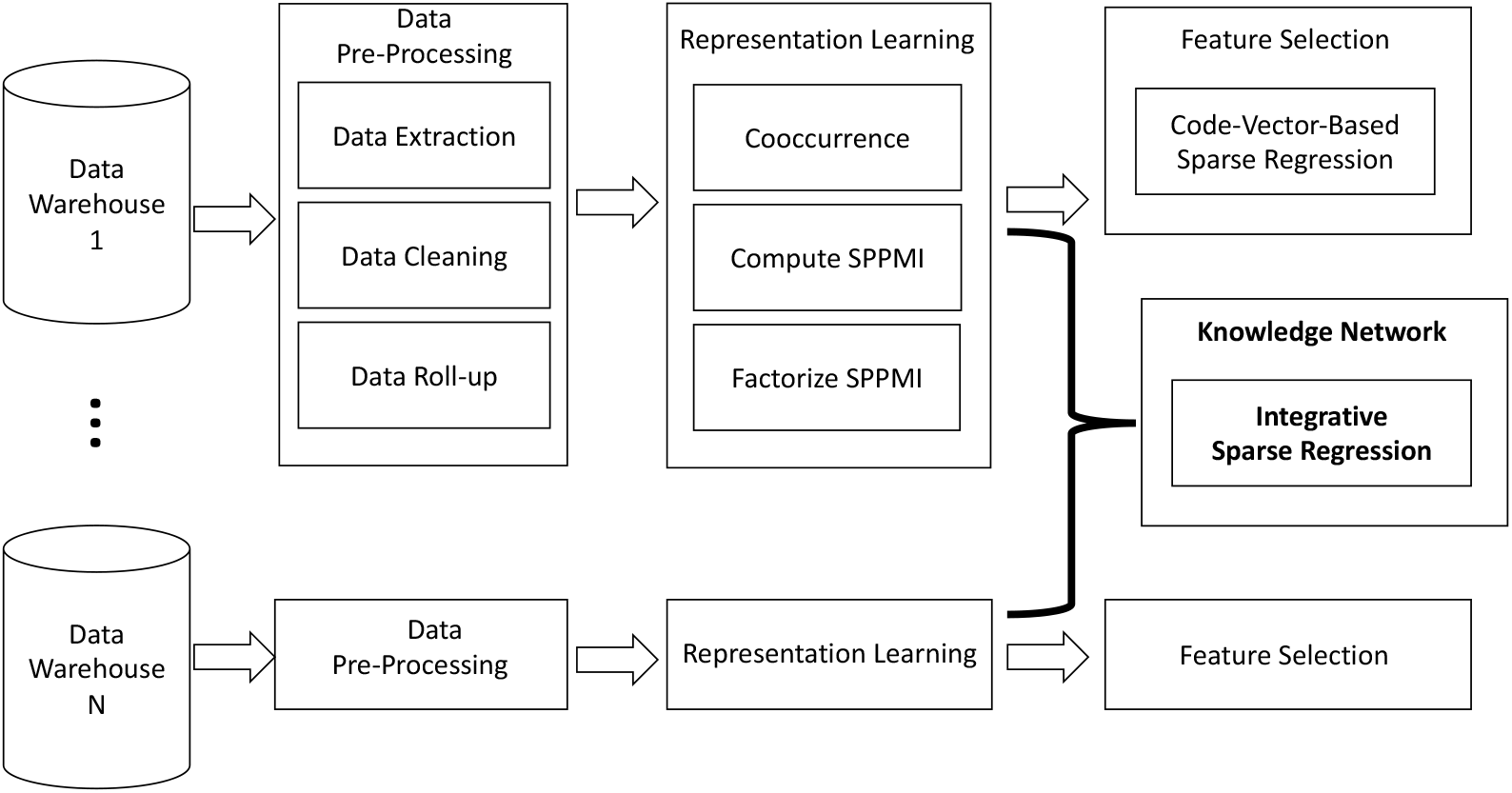
Overview of KESER procedure.

**Figure 2.**
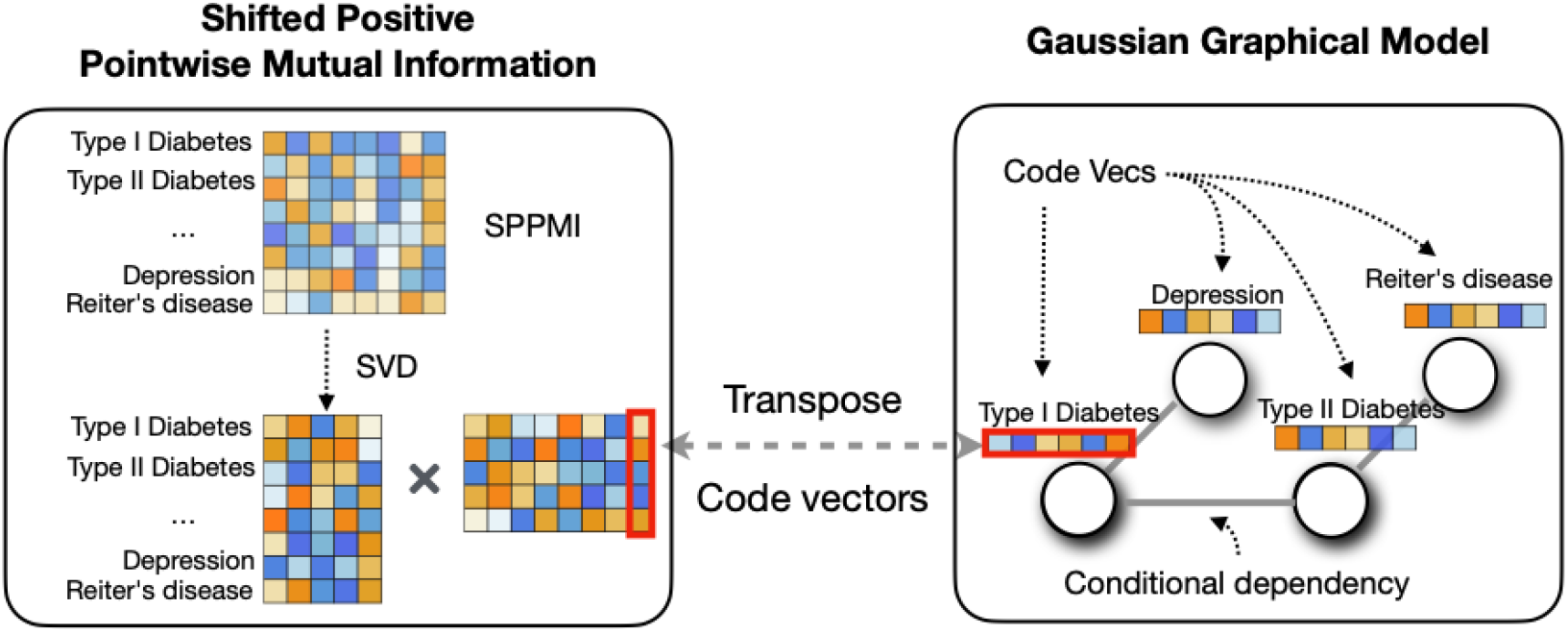
The left panel describes the key steps for learning the embedding vectors: we conduct singular vector decomposition (SVD) on the SPPMI. The right panel describes the statistical model: the embedding vectors follow a Gaussian graphical model where each node of the graph is represented by the vectors.

### Data Pre-Processing

We used EHR data from two large hospital systems, the VA Corporate Data Warehouse (CDW) and Mass General Brigham (MGB). After aggregating four codified data domains (i.e., diagnosis, procedures, lab measurements, and medications) into PheCode, CCS, RxNorm, LOINC codes, and manual lab concepts, and applying frequency control, we finally obtained a total of 9,211 codes at VA and 5,507 at MGB.

### Creating and Validating Embedding Vectors

We obtained embeddings by performing singular value decomposition (SVD) on the shifted positive pointwise mutual information (SPPMI) matrix, known as the SVD-SPPMI algorithm.

#### Known Relation Benchmark relation pairs

To select optimal hyper-parameters and evaluate the performance of the trained embeddings along with the proposed KESER algorithms, we collected a set of known relationship pairs from Wikipedia, PheCode hierarchy, https://www.drugs.com/, MEDRT, SNOMED-CT, and manual annotations. The total numbers of curated relation pairs across all available sources that can be mapped to MGB and VA, as shown in Table S1 of the Supplementary Materials, are 15326 and 15224.

#### Optimal Dimensions of Derived Embeddings

We obtained the initial embedding dimensions by retaining 95% of the variation in the SVD (*d*_*95%*_), resulting in 1800 for MGB and 2900 for VA, as shown in Figure S1 of the Supplementary Materials. We further evaluate strategies for choosing optimal embedding dimensions and the degree to which embedding dimensions may affect the performance of detecting similar concepts and related concepts. We chose the dimensions by maximizing either (a) the signal to noise ratio (SNR); or (b) the area under the receiver operating characteristic curve (AUC) associated with pairs with known relations against random pairs, as detailed in Methods. The dimensions selected to maximize AUC (*d*_*auc*_) tend to be lower than to those selected to maximize SNR (*d*_*snr*_) and selected dimensions are generally lower for assessing similarity compared to those for relatedness. For optimizing similarity assessment, (*d*_*auc*_, *d*_*snr*_) were chosen as (300, 1000) at MGB and (500, 1800) at VA. For detecting relatedness, (*d*_*auc*_, *d*_*snr*_) were chosen as (1800, 1800) at MGB and (2300, 2800) at VA, close to their corresponding *d*_*95%*_.

#### Optimal Window Size and k

We conducted additional sensitivity analyses using different window size and *k* to construct the co-occurrence matrices based on a total of about 70K patients from MGB Biobank. When varying window sizes from 7, 30 up to 60 days and *k* from 1, 5, up to 10, we observed that the embedding quality is the best when *k* = 1 but is not sensitive to the choice of window size (Table S2 of the Supplementary Materials).

#### Performance of Derived Embeddings in Assessing Similarity and Relatedness

Table 1 summarizes the overall accuracy of between-vector cosine similarities in detecting known similarity and relatedness relationships with embedding vectors derived from either SVD-SPPMI or *GloVE*^25^. We focus on *GloVE* trained with dimension 50 and 100 since the *GloVE* algorithm did not converge at higher dimensions. For detecting similar pairs, the SVD-SPPMI based cosine similarities attained an AUC of 0.839 at MGB and 0.888 at VA with dimensions set at *d*_*auc*_. By thresholding cosine similarities to classify pairs as similar with cut-off chosen to maintain false positive rate (FPR) of 0.05 and 0.10, these classifications yielded sensitivities of 0.593 and 0.669 at MGB and 0.679 and 0.772 at VA. For the relatedness, the cosine similarities based on SVD-SPPMI embeddings at *d*_*95%*_ achieved AUC of 0.868 at MGB and 0.862 at VA, sensitivities of 0.608 and 0.717 at MGB and 0.582 and 0.688 at VA at FPR=0.05 and 0.10. Compared to *GloVE*, embeddings derived via SVD-SPPMI achieved similar AUCs but higher sensitivities. As shown in Table S2 of the Supplementary Materials, the accuracy is overall fairly high in assessing most types of relationships including *may cause, differential diagnosis, complications*, and *symptoms* with AUC close to 0.9. The accuracy is lower in detecting *risk factors* and *similar drugs* with AUC close to 0.8. Although assessed using different knowledge sources, these observed levels of accuracy are similar to those previously reported based on embedding vectors trained for natural language processing (NLP) concepts^20^.

**Table 1.** AUCs and sensitivity at FPR = 0.01, 0.05 and 0.10 of between-vector cosine similarity in detecting known similar pairs (RxNorm-RxNorm and Lab-Lab) and related pairs (PheCode-PheCode; PheCode-RxNorm) with embeddings trained via SVD-SPPMI at different choices of dimensions *d*.

| Relation Type | Embedding |  | AUC |  | Sensitivity |  |  |  |  |  |
| --- | --- | --- | --- | --- | --- | --- | --- | --- | --- | --- |
|  |  |  |  |  | FPR=0.01 |  | FPR=0.05 |  | FPR=0.1 |  |
| | Method | $d$ | MGB | VA | MGB | VA | MGB | VA | MGB | VA |
| Similar | GloVe | 50 | 0.869 | 0.860 | 0.425 | 0.386 | 0.603 | 0.620 | 0.686 | 0.704 |
|  |  | 100 | 0.876 | 0.855 | 0.433 | 0.391 | 0.614 | 0.588 | 0.681 | 0.680 |
|  | SVD | 100 | 0.831 | 0.857 | 0.390 | 0.268 | 0.559 | 0.499 | 0.626 | 0.646 |
|  |  | 500 | 0.842 | 0.888 | 0.467 | 0.403 | 0.607 | 0.679 | 0.673 | 0.772 |
| | | $d_{snr}^{(1000,1800)}$ | 0.837 | 0.870 | 0.473 | 0.408 | 0.602 | 0.631 | 0.670 | 0.738 |
| | | $d_{auc}^{(300,500)}$ | 0.839 | 0.888 | 0.455 | 0.403 | 0.593 | 0.679 | 0.669 | 0.772 |
| | | $d_{95\%}^{(1800,2900)}$ | 0.836 | 0.868 | 0.465 | 0.386 | 0.601 | 0.638 | 0.677 | 0.734 |
| Related | GloVe | 50 | 0.873 | 0.805 | 0.275 | 0.198 | 0.538 | 0.384 | 0.659 | 0.505 |
|  |  | 100 | 0.876 | 0.828 | 0.286 | 0.247 | 0.542 | 0.435 | 0.672 | 0.558 |
|  | SVD | 100 | 0.854 | 0.817 | 0.205 | 0.197 | 0.498 | 0.410 | 0.647 | 0.538 |
|  |  | 500 | 0.864 | 0.854 | 0.304 | 0.291 | 0.589 | 0.543 | 0.705 | 0.660 |
| | | $d_{snr}^{(1800,2800)}$ | 0.868 | 0.861 | 0.357 | 0.325 | 0.620 | 0.584 | 0.716 | 0.686 |
| | | $d_{auc}^{(1800,2300)}$ | 0.868 | 0.862 | 0.352 | 0.333 | 0.608 | 0.582 | 0.717 | 0.687 |
| | | $d_{95\%}^{(1800,2900)}$ | 0.868 | 0.862 | 0.352 | 0.333 | 0.608 | 0.582 | 0.717 | 0.688 |

#### Performance of Derived Embeddings in Mapping codes across institutions

Similar to language translation, we learned orthogonal transformation between embedding vectors across the two institutions to enable mapping of a given VA code to the corresponding MGB code ^26^. As summarized in Table S3 of the supplementary Materials, the top-1 and top-5 accuracy of code mapping is around 38% and 67% for VA medication codes → RXNORM and around 42% and 74% for PheCode → PheCode using embeddings of dimension *d*_*auc*_. The code mapping accuracy is fairly comparable when using a larger *d*_*snr*_. The observed code mapping accuracy is comparable to the translation accuracy between different languages reported in the literature ^26 27^.

### Knowledge Extraction via KESER

The KESER approach was developed to select features by using embeddings trained within a specific healthcare center, as well as by leveraging embeddings from multiple healthcare centers while incorporating between-site heterogeneity.

#### Performance of KESER in Detecting Known Relationships

In Table 2, we summarize the average sensitivities and FPR of KESER integrative knowledge extraction using embedding data from both MGB and VA (KESER_INT_) in detecting known associations. For comparison, we also provide results based on KESER performed using MGB data only (KESER_MGB_) and using VA data only (KESER_VA_). The integrative analysis based on KESER_INT_ attained a sensitivity of 0.660 in detecting known related pairs, while maintaining FPR below 5%. The KESER_INT_ algorithm attained accuracy substantially higher than those from KESER algorithms trained with single institution data and the accuracy is generally higher using embeddings from SVD-SPPMI compared to those from *GloVE*.

**Table 2.** Sensitivity and FPR of KESER_MGB_ (MGB), KESER_VA_ (VA) and KESER_INT_ (INT) in detecting known related pairs using embedding vectors trained via GloVE or SVD-SPPMI at various dimensions.

| Embedding Method | Dimension | Sensitivity |  |  | FPR |  |  |
| --- | --- | --- | --- | --- | --- | --- | --- |
|  |  | MGB | VA | INT | MGB | VA | INT |
| GloVe | 100 | 0.399 | 0.356 | 0.438 | 0.021 | 0.030 | 0.038 |
| SVD | 100 | 0.345 | 0.240 | 0.352 | 0.017 | 0.019 | 0.026 |
|  | 500 | 0.453 | 0.368 | 0.526 | 0.021 | 0.022 | 0.035 |
| | $d_{95\%}^{(1800,2900)}$ | 0.531 | 0.489 | 0.628 | 0.027 | 0.027 | 0.042 |

#### Performance of KESER in Identifying Drugs of Rheumatoid Arthritis (RA)

The performance of KESER_INT_, KESER_MGB_, and KESER_VA_ using embeddings obtained by GloVE or SVD-SPPMI in detecting 16 medications commonly used to treat RA is summarized in Table 3. Out of the 16 medications, using embedding from SVD-SPPMI at *d*_*95%*,_ the numbers of drugs selected by KESER_MGB_, KESER_VA_ and KESER_INT_ were 16, 14 and 16, respectively, yielding a sensitivity of 1.00, 0.88 and 1.00. Sensitivity in detecting these medications based on lower dimensional embeddings from SVD-SPPMI or GloVE are generally lower. For example, the sensitivity ranged from 0.41 to 0.53 based on GloVE at d=100 and from 0.82 to 0.94 based on SVD-SPPMI at d=500.

**Table 3.** Sensitivities of KESER_MGB_ (MGB), KESER_VA_ (VA) and KESER_INT_ (INT) in detecting two categories (CAT) of RA related medications. CAT=1 for DMARDs in use and CAT = 2 for other drugs often used to manage RA patients.

|  |  | GloVe |  |  | SVD |  |  |  |  |  |  |  |  |
| --- | --- | --- | --- | --- | --- | --- | --- | --- | --- | --- | --- | --- | --- |
| | | $d = 100$ | | | $d = 100$ | | | $d=500$ | | | $d = d_{95\%}$ | | |
| med | CAT | MGB | VA | INT | MGB | VA | INT | MGB | VA | INT | MGB | VA | INT |
| *abatacept* | 1 | 0 | 0 | 0 | 1 | 0 | 1 | 1 | 1 | 1 | 1 | 1 | 1 |
| *anakinra* | 1 | 0 | 0 | 0 | 0 | 0 | 1 | 1 | 1 | 1 | 1 | 1 | 1 |
| *rituximab* | 1 | 0 | 0 | 0 | 0 | 0 | 0 | 0 | 1 | 1 | 1 | 1 | 1 |
| *tocilizumab* | 1 | 1 | 0 | 1 | 0 | 1 | 1 | 1 | 1 | 1 | 1 | 1 | 1 |
| *tofacitinib* | 1 | 0 | 0 | 0 | 1 | 0 | 1 | 1 | 1 | 1 | 1 | 1 | 1 |
| *adalimumab* | 1 | 0 | 1 | 0 | 1 | 0 | 0 | 1 | 0 | 1 | 1 | 1 | 1 |
| *certolizumab* | 1 | 0 | 0 | 0 | 0 | 0 | 0 | 1 | 0 | 1 | 1 | 1 | 1 |
| *etanercept* | 1 | 1 | 1 | 1 | 0 | 0 | 1 | 1 | 1 | 1 | 1 | 1 | 1 |
| *golimumab* | 1 | 0 | 0 | 0 | 1 | 0 | 0 | 1 | 1 | 1 | 1 | 1 | 1 |
| *infliximab* | 1 | 0 | 0 | 0 | 0 | 1 | 1 | 1 | 1 | 1 | 1 | 1 | 1 |
| *leflunomide* | 1 | 1 | 1 | 1 | 1 | 1 | 1 | 1 | 1 | 1 | 1 | 1 | 1 |
| *hydroxychloroquine* | 1 | 1 | 1 | 1 | 1 | 0 | 1 | 1 | 1 | 1 | 1 | 1 | 1 |
| *sulfasalazine* | 1 | 1 | 1 | 1 | 0 | 0 | 0 | 0 | 1 | 1 | 1 | 1 | 1 |
| *methotrexate* | 1 | 1 | 1 | 1 | 0 | 1 | 1 | 1 | 1 | 1 | 1 | 1 | 1 |
| *methylprednisolone* | 2 | 1 | 1 | 1 | 0 | 0 | 1 | 0 | 0 | 0 | 1 | 0 | 1 |
| *prednisone* | 2 | 0 | 1 | 0 | 0 | 0 | 0 | 1 | 1 | 1 | 1 | 1 | 1 |
| *folic acid* | 2 | 1 | 1 | 0 | 1 | 0 | 1 | 1 | 1 | 1 | 1 | 0 | 1 |
| **Sensitivity** |  | **0.471** | **0.529** | **0.412** | **0.412** | **0.235** | **0.647** | **0.824** | **0.824** | **0.941** | **1** | **0.882** | **1** |

#### Validation of KESER using Real World EHR Phenotyping

We conducted KESER feature selection for 8 diseases: coronary artery disease (CAD), type I diabetes mellitus (T1DM), type II diabetes mellitus (T2DM), depression, rheumatoid arthritis (RA), multiple sclerosis (MS), Crohn’s disease (CD) and ulcerative colitis (UC). Figure 3 shows KESER-selected features for RA and UC. Results for the remaining six diseases are summarized in Figures S2-S9 in the Supplementary Materials. Since the goal of the feature selection is to achieve high sensitivity, i.e., to identify many of the potentially important features, less emphasis should be placed on the magnitude of the sparse regression coefficients. The results were largely consistent with clinical knowledge. For RA, the five most important codes were *tofacitinib, tocilizumab, golimumab, abatacept and methotrexate*, all current therapies for RA. Other selected features include differential diagnoses for RA (e.g. *juvenile rheumatoid arthritis, osteoporosis, psoriasis*) and lab tests for diagnosing or monitoring RA (e.g. *cyclic citrullinated peptide, c-reactive protein and erythrocyte sedimentation rate*). Inflammatory bowel disease (IBD) comprises two subtypes, CD and UC. For UC, top features selected by KESER consisted of treatments currently used to treat the condition. While *vedolizumab* is used in both UC and CD, *golimumab* is indicated for UC and not CD (Figure S9). UC features also include CD and *noninfectious gasteroenteritis* as differential diagnoses as well as important procedures such as colonoscopy, proctoscopy and colorectal resection.

**Figure 3.**
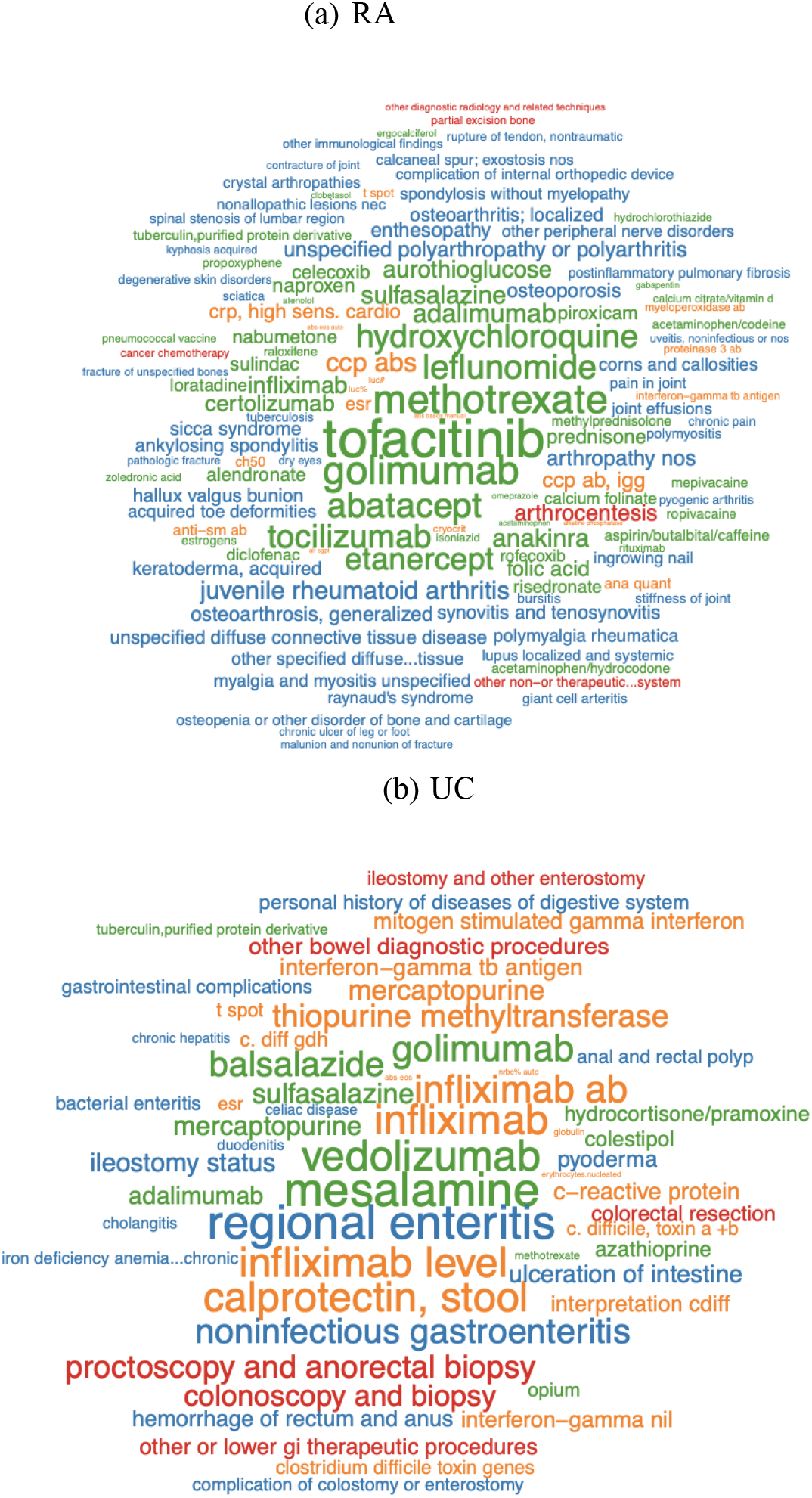
Word cloud for KESER_MGB_ selected features of Rheumatoid Arthritis (RA) and Ulcerative Colitis (UC). The size of the words are proportional to the absolute coefficients from the embedding regression.

Using codified EHR data from 68,213 MGB Biobank participants, we compared the performance of two supervised phenotype algorithms, the adaptive LASSO (aLASSO) and random forest (RF), trained with existing feature selection strategies to those trained with KESER-selected features. Those existing feature selection strategies included the main PheCode of the disease only (PheCode), all features (FULL), or informative features selected manually or extracted using unsupervised algorithms such as SAFE^15^. The accuracies of the aLASSO phenotyping algorithms trained with different feature sets are summarized in Figure 4 and more detailed comparisons including the RF results are given in Figure S10 of the Supplementary Materials. Given the same feature set, the RF algorithms generally performed slightly worse than the aLASSO algorithms in part due to overfitting. The relative performance of the RF algorithms trained with different feature sets is similar to those from aLASSO. The algorithms generally attained higher performance using embeddings from SVD-SPPMI than those from GloVE. The results are quite similar when using KESER_INT_ versus KESER_MGB_ and hence using MGB embedding information may be sufficient for phenotyping at MGB. Hence we focus our discussions below on the aLASSO algorithms and for KESER, we focus on KESER_MGB_ with SVD-SPPMI embeddings for brevity. Across the 8 phenotypes, phenotyping algorithms trained via aLASSO with KESER_MGB_-selected features attained higher AUCs and F-scores than those based on PheCode alone or using FULL features, and similar AUCs as those trained with SAFE features. On average, the AUC of KESER_MGB_ with SVD-SPPMI based algorithms was 0.052, 0.144 and 0.007 higher than those based on PheCode, FULL and SAFE features. The average F-score of KESER_MGB_ based algorithms was 0.173, 0.157 and 0.013 higher than those based on PheCode, FULL and SAFE features. The 95% confidence intervals of the accuracies associated with algorithms trained with KESER-selected features are similar to the SAFE features, while those of the FULL features and main PheCode alone are substantially wider.

**Figure 4.**
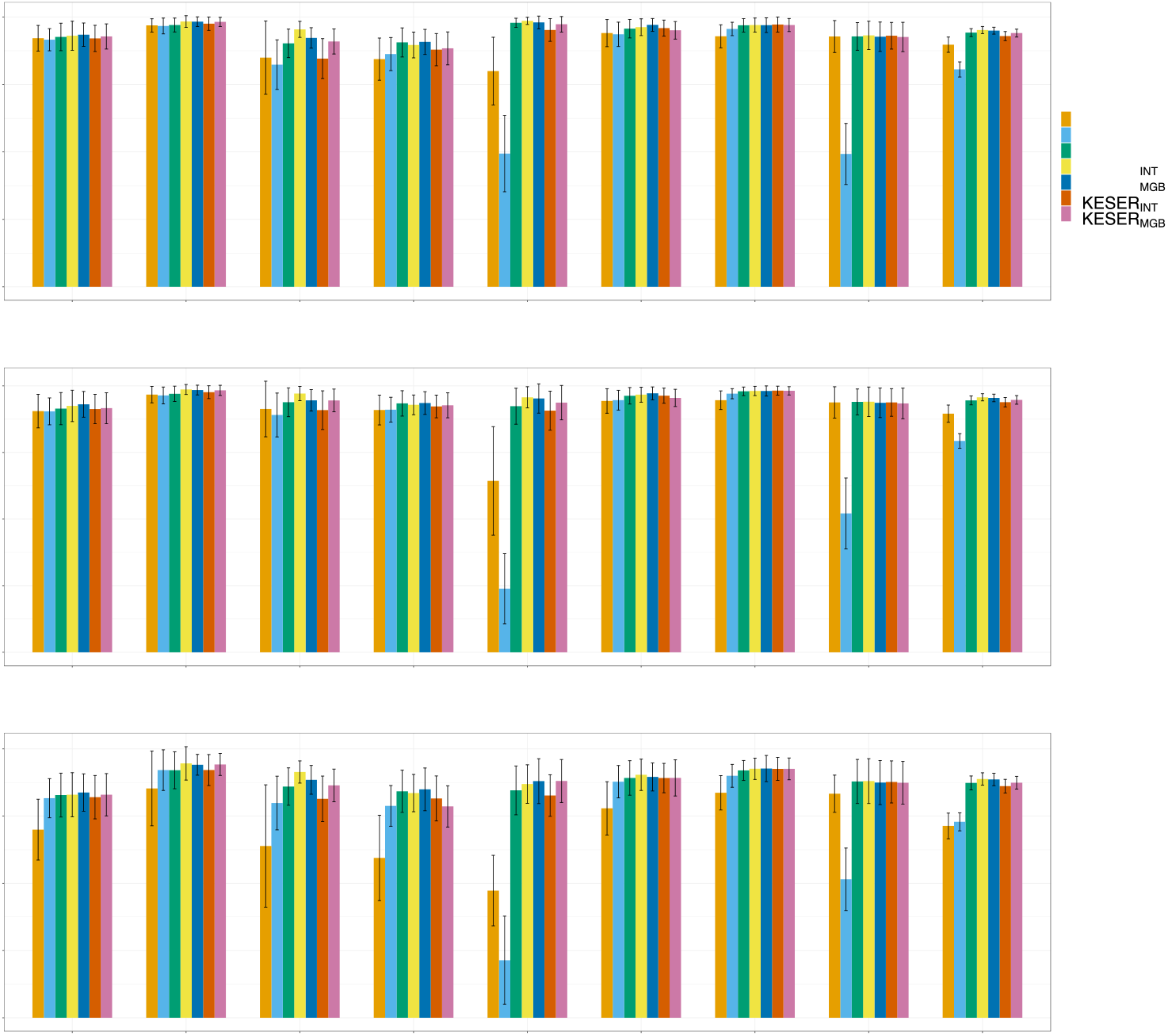
Comparison of AUCROCs, AUCPRCs and F-scores with gold standard labels for adaptive lasso phenotyping algorithms for 8 diseases using the main PheCode only (PheCode), all features (FULL), SAFE selected features (SAFE), KESER_MGB_ and KESER_INT_ selected features based on SVD-SPPMI embeddings as well as KESER_MGB_ and KESER_INT_ selected features based on GloVE embeddings. F-scores are calculated at the cutoff points with the estimated prevalence equal to the population prevalence. The bootstrap based 95% confidence intervals (bars) are shown.

### Knowledge Mapping by Performing Node-wise KESER

We summarized the clinical knowledge network, namely a *knowledge mapping*, by performing node-wise KESER across all PheCode and RxNorm (https://github.com/celehs/KESER). Figure 5 is a screenshot of the webAPI, given a specific target drug, RxNorm 214555 for *etanercept*. The node-wise knowledge extraction aims to find the neighborhood codes related to the target code *etanercept*. Figure 5(A) displays codes connected to *etanercept* from KESER_INT_, which consists of 36 PheCodes, 49 RxNorm codes, 3 CCS codes and 15 lab codes. Confirmed by domain experts, the results were largely consistent with clinical knowledge. For example, diseases commonly treated by *etanercept*, such as sacroiliitis, juvenile rheumatoid arthritis, RA, ankylosing spondylitis, were selected by the knowledge network. Drugs, procedures and lab tests usually used together with *etanercept*, such as *methotrexate, arthrocentesis, HLAB27*, and *CRP*, were also selected. Figures 5(B) and 5(C) display the local network based on KESER_VA_ and KESER_MGB_. We observed four lab codes uniquely identified by VA and nine lab codes uniquely identified by MGB. The discrepancy of the local networks at VA and RPDR lies only in lab codes. This is expected because the majority of the lab codes are unique to the site, resulting in high cross-site heterogeneity in lab coding. By integrating data from both sites, KESER_INT_ is able to achieve higher accuracy in reflecting clinical knowledge.

**Figure 5.**
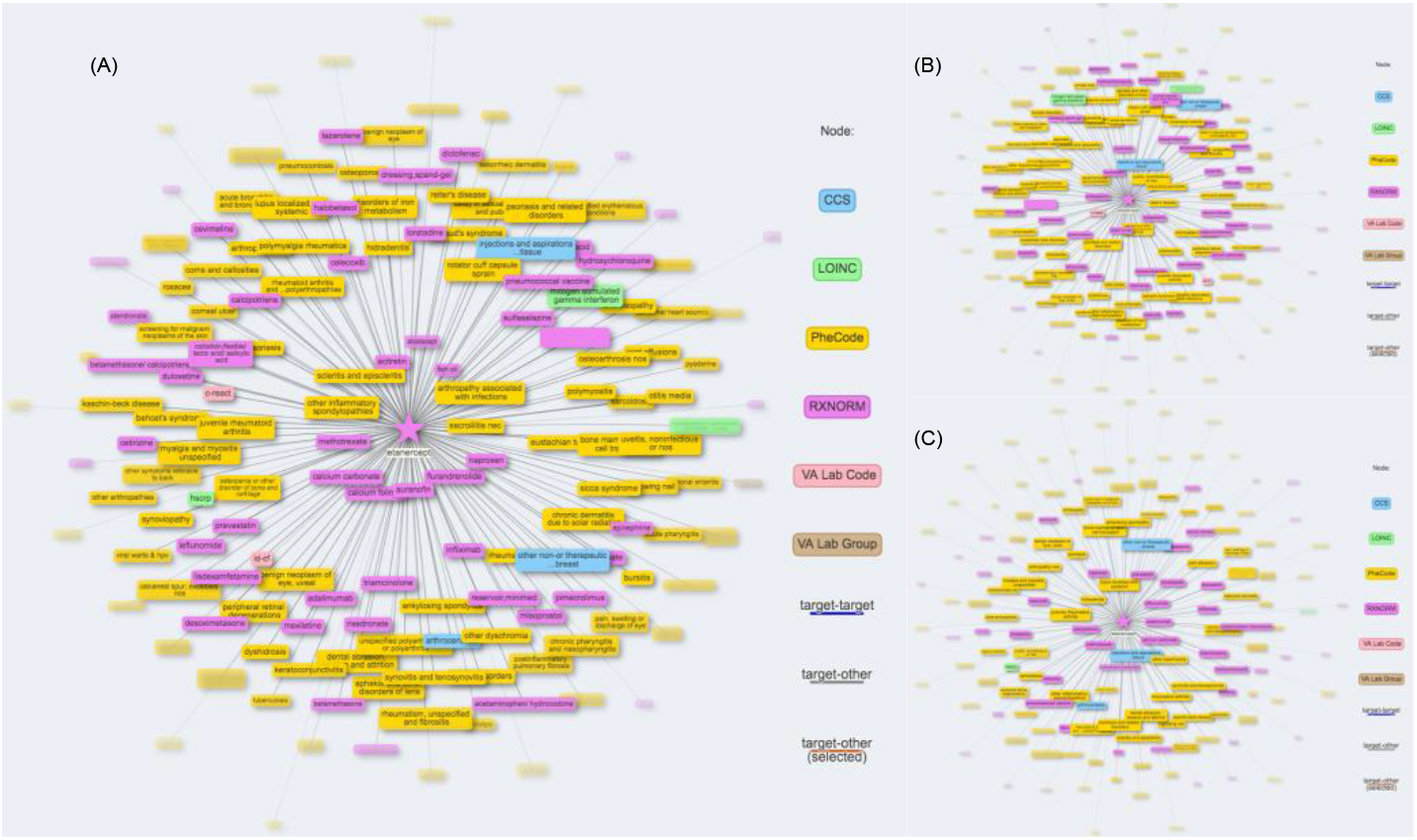
Clinical knowledge network for Etanercept learned based on (A) KESER_INT_; (B) KESER_VA_; (C) KESER_MGB_.

These results demonstrate that KESER can successfully select informative and clinically meaningful features that can be used effectively for phenotyping and other downstream analyses.

## DISCUSSION

The KESER approach efficiently summarizes patient-level longitudinal EHR data into hospital-specific embedding data and enables extraction of clinical knowledge based only on summary level data. This summary data generated based solely on relationships between codes, and clusters related codes together, which provides ready information on features that may be important for identifying or studying different phenotypes. The KESER approach enables assessment of conditional dependency between EHR features by performing sparse regression of embedding vectors without requiring additional patient level data. In this paper we demonstrate the advantage of integrative analyses across sites in detecting known associations. Ultimately, we believe this innovation provides a potential solution for barriers facing the much-needed multi-center collaborative studies using EHR data.

The majority of EHR-based clinical studies are performed entirely behind the firewalls of individual institutions. Collaborations across centers typically require that each institution perform analyses individually with results compared across institutions. However, coding behaviors, disease management and strategies, and healthcare delivery patterns^28^ can vary across different healthcare systems. For example, at VA, medication procedures (such as infliximab-injection) are coded as HCPCS procedure codes, while at MGB, they are coded as local medication codes that directly map to RxNorm. At VA, the majority of patients are male, and thus the pattern of diseases or treatments may differ from MGB where females are the majority. Variations between the two institutions were observed when validating the embedding vectors compared against known PheCode-RxNorm pairs (Table 1). While the knowledge derived from the embedding vectors captures all the relevant RA treatments at both VA and MGB, the weights of the individual treatments differed slightly between the two healthcare systems (Figure 3). Among the top-50 weighted treatments, there are 36 same concepts obtained from both healthcare systems. At VA *methotrexate* had the largest coefficient compared to *tofacinitib* at MGB. Integrating the data from both systems improves the robustness of the identified relationships and accounts for the heterogeneity of data in each system. Notably, since the embedding vectors contain no patient data, the integration of these data can be performed outside of each system.

Embedding vectors also provide information on highly related groups of codes. Unlike ICD codes which have established groupings and hierarchies, lab codes are much less standardized, and no established grouping structure can be used at scale for research studies. As an example, for the inflammatory marker C-reactive protein (CRP), potential lab codes include, LOINC:11039-5 (crp), LOINC:30522-7 (crp, high sens, cardio), and LOINC:X1166-8 (crp (mg/L)). Additionally, at both VA and MGB, individual labs within each institution also had unique lab codes that do not map to the LOINC codes. The embedding vectors derived from the co-occurrence matrices enable grouping of codes based on the similarity between the vectors, thus allowing the use of grouped lab codes in research studies.

We also addressed the need to tailor the dimension of embedding vectors to the goals of a particular study. Currently, there is no clear evidence regarding how to select the optimal dimension for analyses using embeddings. Existing embedding-based approaches usually use a 300-dimension word embedding *GloVE*^25^ or a 500-dimension CUI embedding for *cui2vec*^20^. We demonstrate that different dimensions may be preferred for different tasks. Lower dimensions appear to be better suited for the task of identifying near synonymous concepts or translations while higher dimensions are needed for assessing relatedness and embedding regression aiming to optimize feature selection and building knowledge networks. While lower dimensions may be useful for many downstream tasks such as code mapping between institutions, we recommend keeping embedding vectors at high dimensions for dissemination to enable better assessment of relatedness while allowing users to further truncate to lower dimensions for other tasks.

In this paper, we derived embeddings via SVD-SPPMI, considered in the literature as equivalent to the skip-gram algorithm with negative sampling (SGNS)^17^. Computationally, the SVD-SPPMI approach is substantially more efficient than SGNS as it does not need to conduct the negative sampling which is computationally intensive especially when the number of codes is massive. Due to both IRB and computational constraints, we are only able to derive embeddings from SVD-SPPMI and *GloVE* which only require summary data and but not SGNS. We find that SVD-SPPMI derived embeddings generally have more robust performance compared to those from GloVE which also appears to suffer from convergence issues when fitting for higher dimensions, possibly due to the sparsity of the SPPMI matrices.

The embedding vectors provide not only a method to share and analyze data, but also an opportunity to develop an integrated clinical knowledge network with input from many institutions. This network allows us to visualize the node-wise relationships between a target code (e.g., a PheCode or a RxNorm) and its neighborhood codes: PheCode, RxNorm, CCS and Labs (Figure 5). By leveraging information from both sites, the integrative network covers all available knowledge and consists of a more comprehensive pool of neighborhood codes compared with local networks.

Finally, using KESER, this knowledge network can be updated over time to study relationships between emerging conditions and their relationships with existing conditions, across multiple healthcare systems. This is particularly relevant for future studies on the impact of the COVID-19 pandemic. There is still a lack of knowledge in fundamental aspects of COVID-19, such as the development, management and treatment of the disease, and how those aspects differ across different sites and countries. Therefore, creating an integrated clinical knowledge map of codified data for COVID-19 will be of great interest. This knowledge map can be then used to facilitate the classification of COVID-19 patients with selected features. As an exploratory analysis, we constructed two separate co-occurrence matrices and derived embeddings via the SVD-SPPMI using all EHR data up to Nov, 2020 from 30K COVID+ patients at MGB and 100K COVID+ patients at VA. As a proof of concept, we identified clinical concepts most related to the COVID code. As shown in Figure S11 of the Supplementary Materials, the results are encouraging in that the top selected codes include the highly important laboratory tests for monitoring COVID progression (e.g. *D-dimer, CRP, Ferritin*) and medications for managing COVID patients (e.g. *norepinephrine* often used as first line vasoactive, *cefepime* for managing bacteria pneumonia complications, *tocilizumab, dexamethasone* and *remdesivir*) as well as related diagnoses and complications (e.g. *viral pneumonia, respiratory insufficiency, shock*, and *kawasaki disease*).

In conclusion, KESER provides an approach allowing investigators to integrate patient level data as embedding vectors from multiple EHR systems for downstream analyses. We provide an example of using the knowledge network to automatically provide features that may be important for phenotyping, without requiring additional patient level data. This innovation will facilitate multi-center collaborations and bring the field closer to the promise of creating distributed networks for learning across institutions while maintaining patient privacy.

## METHODS

We highlight three key innovations detailed below in the methods. First, we provided an approach to integrate four domains of codified data, ICD, CPT, laboratory codes, and medications, from two large hospital systems. Second, we applied a data driven approach to specify the dimension of embedding vectors. Third, we developed a method to use embedding vectors rather than patient-level data as the input into a sparse graphical model.

### Data Pre-Processing

#### Data Sources

##### The VA Corporate Data Warehouse (CDW)

aggregates EHR data from over 150 VA facilities into a single data warehouse. It contains clinical, financial, and administrative records for over 23 million unique individuals (1999-2019). The CDW supports both business operations and research. A total of 12.6 million patients with inpatient and outpatient codified data from at least 1 visit were included for this analysis. We defined outpatient visit to include services from all VA outpatient stop codes. There are over 500 outpatient stop codes that cover a wide range of services such as emergency department visits, therapy and primary care. We first extracted records from the CDW. We then grouped each patient’s records together in ascending chronological order. Codes occurring multiple times for the same patient within the same day are counted once per day. The resulting files were stored using parquet, a columnar storage format. The parquet file format was well suited to storing this data compactly while also allowing parallel processing.

##### Mass General Brigham (MGB), formerly Partners Healthcare

is a Boston-based non-profit healthcare system anchored by two tertiary care centers, Brigham and Women’s Hospital (BWH) and Massachusetts General Hospital (MGH). The Research Patient Data Registry (RPDR) of MGB is a research copy of the electronic health records of BWH and MGH with over 1 billion visits containing diagnoses, medications, procedures, and laboratories information. The patient population included 2.5 million patients with at least 3 visits spanning more than 30 days. The analysis included coded data from all inpatient, outpatient and emergency department visits between 1998 and 2018. We used the same format as VA described above to store patient visit level data for processing.

#### Code Roll-up

We gathered four domains of codified data including diagnosis, procedures, lab measurements, and medications from VA and MGB EHRs. Since multiple EHR codes can represent the same broad concept, (e.g. acute myocardial infarction (MI) of anterolateral wall and acute MI of inferolateral wall are separate codes that describe the same concept of MI), we rolled individual codes to a code representing a general concept. ICD codes were aggregated into PheCodes to represent more general diagnoses, e.g., MI rather than acute MI of inferolateral wall, using the ICD-to-PheCode mapping from PheWAS catalog (https://phewascatalog.org/phecodes). We utilized multiple levels of granularity of PheCode, including integer level, one-digit level and two-digit level. To reduce the effect of collinearity, when conducting KESER regression, for phenotypes with multiple levels of PheCode, we only included one-digit level PheCodes.

For procedure codes, including CPT-4, HCPCS, ICD-9-PCS, ICD-10-PCS (except for medication procedures), we assigned CCS categories based on *the clinical classification software (CCS) mapping* (https://www.hcup-us.ahrq.gov/toolssoftware/ccs_svcsproc/ccssvcproc.jsp). For medication codes, we aggregated the local medication codes at VA and MGB to the ingredient level RxNorm codes^29^.

For laboratory measurements, due to the difference in coding systems between VA and MGB, we created a code dictionary for each site. At VA this was done by grouping local lab codes to manually annotated lab concepts or LOINC codes, as well as individual lab codes that have not been annotated but occurred in at least 1000 patients. At MGB, all local lab codes were aggregated into group and a LOINC code was assigned to each. Since embeddings cannot be trained well for very low frequency codes, we only included codes occurring >1000 times at MGB and >5000 times at VA. The different thresholds were used because VA has a larger population and larger number of codes than MGB. A total of 9,535 codes (1776 PheCodes, 1561 RxNorms, 5974 Labs and 224 CCS groups) at VA and 5,245 codes (1772 PheCodes, 1238 RxNorms, 1992 Labs and 243 CCS groups) at MGB passed the frequency control.

### Creating Embedding Vectors

We obtained embeddings by performing singular value decomposition (SVD) on the shifted positive pointwise mutual information (SPPMI) matrix, known as the SPPMI-SVD algorithm. This approach provided embeddings considered as efficient and equivalent to those derived from the skip-gram algorithm with negative sampling^17,20,30,31^.

#### Co-occurrence Matrix

We first constructed code co-occurrence matrices as described in Beam et al^14^. For any given patient, we scanned through each of their codes as a target code. For any given target code occurring at time *t*, denoted by *w*_*t*_, we counted all codes occurring within 30 days of *t* as co-occurrences with *w*_*t*_. The total numbers of co-occurrences for all possible pairs of codes are aggregated over all target codes within each patient and then across all patients, yielding the co-occurrence matrix, denoted by ℂ = [C(*w, c*)]. Although only codes that occur after the target code are considered, this is the same as finding co-occurring codes within 30 days of the target code (i.e. between −30 and 30 days), owing to the symmetry of the data. Thus, given a target phenotype *w* (e.g, PheCode 714.1 for RA), we assume the context codes vocabulary 𝒱_*c*_ (*w*) are the codes co-occurred with the target word within a 30-day window. This step requires considerable computational resources and a detailed algorithm for efficiently computing the co-occurrence matrix was created for this study (https://github.com/rusheniii/LargeScaleClinicalEmbedding).

Since our sparse regression procedures (described in later sections) require selection of tuning parameters, we constructed two separate co-occurrence matrices at each site. At VA, from the 12.6 million patients, we used data from 11.6 million patients to create a training matrix 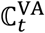 and data from the remaining 1 million patients to create a validation matrix 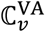. At MGB, we used half of the patients to create training and the other half to create validation matrices, respectively denoted by 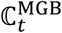 and 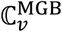.

#### Embedding via SVD of the SPPMI matrix

We calculate the SPPMI matrix as:

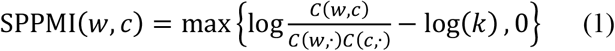

with the negative sample *k* set as 1 (i.e. no shifting), where *c*(*w*,.) is the row sum of *c*(*w, c*). For each given SPPMI, we obtain its first *d*-dimensional SVD as 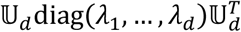 and then construct the *d*-dimensional embedding vectors as 𝕍_*d*_, where 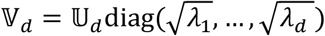.

#### Knowledge Extraction via Sparse Embedding Regression (KESER)

We propose to infer conditional dependency among the clinical codes based on the conditional dependency among their corresponding embedding vectors. To provide a rationale for this framework, we note that the skip-gram model with negative sampling^16^ directly encodes the *marginal* dependency between the target code *w* and its context code *c* via the covariance between their respective embedding vectors ***V***_*w*_ and ***V***_*c*_ with

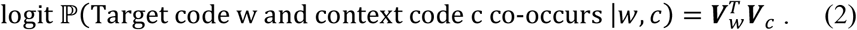

This motivates us to model the *conditional* distribution of the target code *w* and other codes by imposing a Gaussian distribution on the embedding vectors and inferring the dependency via a Gaussian graphical model on top of the skip-gram model. Specifically, in the *m*^*th*^ healthcare center, we assume that the embedding vector of code 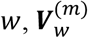, follows a conditional vector-valued Gaussian distribution centered at the linear combination of its context word vectors, i.e.,

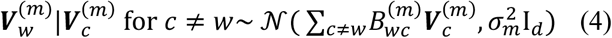

for *m* = 1, … *M*, where 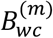 encodes the conditional dependency between codes *w* and *c*, with 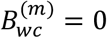 if 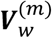 is independent of 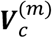 given all other code vectors. For symmetry, we assume 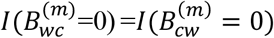 for all target words *w* and the context *c*. Figure 2 visualizes the two-layer hierarchical structure of our model. The sparsity structure of 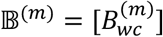 enables us to infer about the relatedness among different features for the *m*^*th*^ healthcare center, which can be used for both feature selection and learning a knowledge graph.

#### Site-Level Feature Selection

Due to the heterogeneity of the coding patterns across healthcare centers, feature selection can be done using embeddings trained within a specific healthcare center. For the *m*^*th*^ center, we select features important for a specific target phenotype *w* (e.g, PheCode 714.1 for RA) by performing an elastic net penalized regression^32^ of 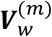 against 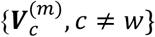. We first perform an initial screening based on marginal cosine similarity and consider codes in 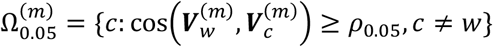 for further selection, where ρ_0.05_ is the upper 5^th^ percentile of the cosine similarity among randomly selected pairs. Since the cosine similarity distribution varies across different relationship types (e.g. PheCode-PheCode versus PheCode-RXNORM), we recommend choosing ρ_0.05_ within each relationship types. Then we estimate 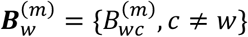 as

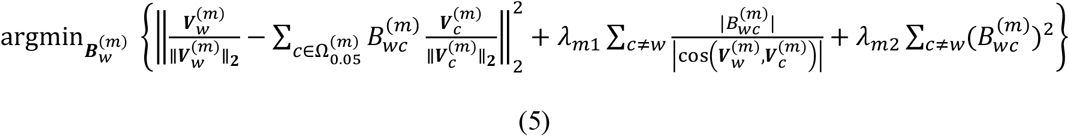

for some tuning parameters λ_*m*1_,λ_*m*2_ > 0 to be selected. Features with 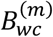 estimated as non-zero are deemed as important for the phenotype *w* in the *m*^*th*^ healthcare center. These selected features can be used for downstream analysis such as developing phenotyping algorithms for a target phenotype. See Appendix A of the Supplementary Materials for details on the tuning of λ_*m*1_ and λ_*m*2_.

#### Integrative Knowledge Network

To learn a knowledge network that encodes relatedness among diseases, procedures, medications, and laboratory tests, we propose to leverage embedding data from multiple healthcare centers while incorporating between-site heterogeneity. Specifically, the conditional dependency structure, as measured by the support of 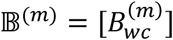 in model (4), are similar across healthcare centers, although the magnitude may differ. Since not all codes are present in all centers, we set 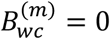 for all 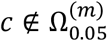. Our goal is to identify the support 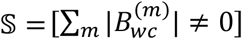 via an integrative analysis of the *M* sets of embedding data 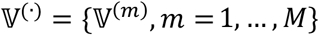. Specifically, for each *w*, we estimate 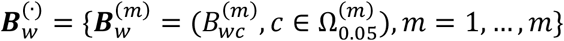 via an integrative least squared regression with a mixture of ridge and group sparse penalty as

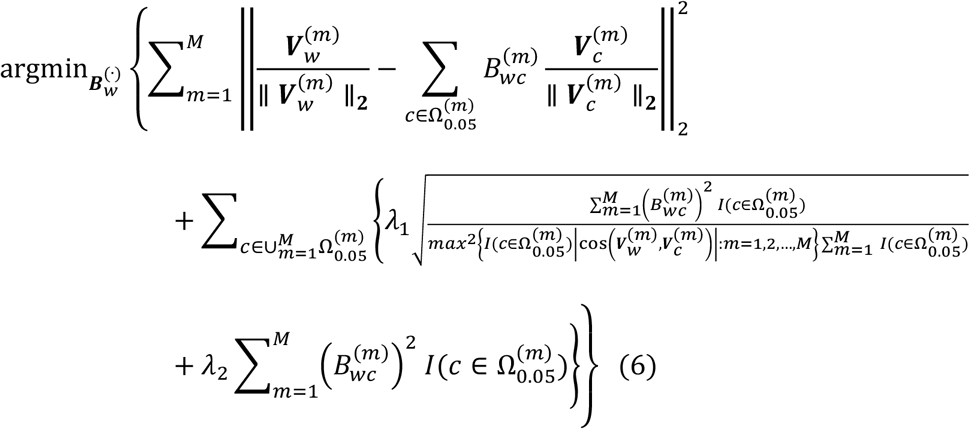

where λ_1_ and λ_2_ are two tuning parameters. Through the group lasso penalty, we are able to borrow signals from all *M* systems and select the important features that appear in multiple sites more efficiently compared with site-specific sparse regression. A complete knowledge network can be established by performing node-wise integrative analysis for each code. See Appendix A of the Supplementary Materials for details on the tuning of λ_1_ and λ_2_.

Tuning parameter selection in this setting differs from standard regression in that the *d*-dimensional embedding vectors are not *d* independent realizations of random variables and hence it is not appropriate to perform cross-validation directly over the embedding vectors. We instead constructed embedding vectors using a training SPPMI matrix and a validation SPPMI matrix, trained with non-overlapping patients, within each healthcare system as described above. See Appendix A of the supplementary materials for a detailed description of the implementation of both KESER feature selection and construction of knowledge network. Source code for implementation can be found at https://github.com/celehs/KESER.

### Evaluation and Validation

#### Evaluation with Known Relation Pairs

To tune hyperparameters and evaluate the performance of the trained embeddings as well as KESER algorithms, we collected a set of known disease-disease (PheCode-PheCode) pairs from Wikipedia and PheCode hierarchy, disease-drug (PheCode-RxNorm) pairs from https://www.drugs.com/ and MEDRT, drug-drug (RxNorm-RxNorm) pairs from SNOMED-CT, and lab-lab pairs from manual annotation. We performed named entity recognition^33^ on the entity pairs extracted from the knowledge sources and mapped these pairs to the text strings of the codified concepts from MGB and VA. Only a small fraction of the extracted known relationship pairs can be mapped directly to the EHR codified concepts due to their difference in encoding and representation.

#### Evaluation of Hyper-parameters

There are several hyper-parameters that may impact the quality of the embeddings including embedding dimension *d*, window size, and shifting parameter *k*. Due to computational constraints, we performed sensitivity analyses to evaluate how window size and *k* impact the embedding quality using the MGB Biobank consisting of EHR data from about 70K patients. We derived embeddings with co-occurrence matrices constructed with window size ranging from 7, 30 up to 60-days, and *k* ranging from 1, 5, to 10. To select the dimension *d*, we first initialized the dimensions by retaining 95% of the variation in the SVD, denoted by *d*_*95%*_. Subsequently, we considered two data-driven strategies for optimizing the dimension up to *d*_*95%*_ by maximizing (i) the signal to nose ratio (SNR); and (ii) the AUC, where SNR(*d*) = *W*_*d*_/*S*_*d*_, *W*_*d*_ and *S*_*d*_ are the average cosine similarity among all pairs with known relationships and among all random pairs. For similarity, we used the PheCode hierarchy for tuning optimal dimensions and defined pairs as similar if they shared the same integer to calculate the SNR and AUC. For relatedness, we used 10% of the known related PheCode-PheCode pairs from Wikipedia and PheCode-RxNorm pairs from https://www.drugs.com/ and MEDRT to tune the dimension and used the remaining known related pairs for validation.

#### Performance of Derived Embeddings and KESER in Detecting Known Relationships

We evaluated the quality of the derived embedding vectors by quantifying their accuracy in detecting known similar pairs (RxNorm-RxNorm and Lab-Lab) and related pairs (PheCode-PheCode, PheCode-RxNorm), and evaluated the KESER algorithm by quantifying its power in detecting known related pairs as described above. For each type of relation, since a vast majority of pairs are unrelated, we randomly sampled a large number of pairs within each type of relationships to obtain the reference distribution for unrelated pairs. For each type of relationship, we obtained the cosine similarity of the embedding vectors between known pairs and between random pairs. We first calculated the area under the AUC as an overall accuracy summary. We then reported the sensitivity of detecting related pairs by thresholding cosine similarities to achieve a false positive rate (FPR) of 0.01, 0.05 or 0.10. We also evaluated the performance of the KESER for feature selection at each site and integrative feature selection at both sites. We report the sensitivities in detecting known related PheCode-PheCode and PheCode-RxNorm pairs, that is the proportion of pairs detected by KESER among all known pairs.

#### Performance of Derived Embeddings in Cross-institution Code Mapping

The trained embeddings at MGB and VA can be used to map codes across the two institutions via orthogonal transformation similar to language translation^26^. Specifically, let 𝕍^(VA)^ and 𝕍^(MGB)^ denote embedding vectors for codes that are common to both institutions. We may find an orthogonal matrix. to minimize the distance between 𝕍^(MGB)^ and 𝕍^(VA)^. as in Smith et al (2017)^26^. We used 1823 codes (223 CCS, 178 LOINC, 698 PheCode and 724 RXNORM) that are common to MGB and VA to train.. The test set consists of 1000 PheCodes that are common to both institutions but not included in the training set as well as a set of manually curated 251 VA local medication code → RXNORM mappings. We evaluate the quality of the cross-institution mapping based on the top-1, top-5 and top-10 accuracy calculated based on the test set. We performed the code-mapping with embeddings of dimensions chosen both via AUC and SNR.

#### Performance of KESER in Identifying of Drugs of Rheumatoid Arthritis (RA)

Patients with RA are treated with disease modifying anti-rheumatic drugs (DMARDs), treatments that can prevent progression of RA. A list of 16 RA treatments approved prior to 2017 were manually curated by domain experts and grouped into two categories: 1) DMARDs currently in use, 2) RA-related drugs used in conjunction with DMARDs. We reported sensitivities in detecting the RA-related drugs using KESER against this manually curated list.

#### Performance of KESER in a Real World EHR Phenotyping Research Application

One downstream application of feature selection is to develop supervised phenotyping algorithms for classifying disease status with these selected features. Supervised algorithms are typically developed using a training dataset consisting of gold standard labels and observations on a given set of candidate features^34^. Existing phenotyping algorithms have considered various approaches to selecting candidate features including the main PheCode of the disease only (PheCode), all features (FULL), or informative features selected manually or extracted using unsupervised algorithms such as SAFE^15^. Using codified EHR data from 68,213 MGB Biobank participants, we compared the performance of supervised phenotype algorithms trained with these existing feature selection strategies to those trained with KESER-selected features. We trained and validated phenotyping algorithms for 8 phenotypes: coronary artery disease (CAD), type I diabetes mellitus (T1DM), type II diabetes mellitus (T2DM), depression, rheumatoid arthritis (RA), multiple sclerosis (MS), Crohn’s disease (CD) and ulcerative colitis (UC), based on gold standard labels manually curated on an average of 545 patients for each disease. For each phenotype, the labeled set was randomly sampled from a filter positive set consisting of patients with at least one relevant PheCode. We corrected for overfitting via.632 bootstrap, a smoothed version of cross validation^35^.

All phenotyping algorithms were trained by fitting adaptive LASSO penalized logistic regression models and random forest models and validated on the subset of labeled patients with at least one PheCode for each disease. We evaluated the accuracy of the phenotyping algorithms based on their area under the receiver operating characteristic curve (AUCROC), the area under the precision-recall curve (AUCPRC) as well as the F-score of the corresponding binary classifiers with threshold values set such that the percentage of patients classified as positive matches the disease prevalence. In addition, we obtained the confidence interval by bootstrap resampling. The phenotyping algorithms were only trained and validated in the filter positive set, since the negative predictive values of the filters are nearly 100% ^36^.

## Data Availability

DATA AVAILABILITY 
The knowledge network is available at https://celehs.hms.harvard.edu/network/

https://celehs.hms.harvard.edu/network/

## ETHICS

The study protocol was approved by the MGB Human Research Committee (IRB00010756). No patient contact occurred in this study which relied on secondary use of data allowing for waiver of informed consent as detailed by 45 CFR 46.116.

These activities were approved through the VA Central IRB. They were supported by Million Veteran Program, VA Central IRB 10-02, and approved under VA Central IRB protocol 18-38.

## DATA AVAILABILITY

The knowledge network is available at https://celehs.hms.harvard.edu/network/.

## COMPETING INTERESTS

The authors declare that there are no competing interests.

## AUTHOR CONTRIBUTION

**Conceptualization**: Tianxi Cai, Katherine P. Liao; **Methodology**: Chuan Hong, Everett Rush, Molei Liu, Doudou Zhou, Ronald Hauser, J. Michael Gaziano, Shawn N. Murphy, George Ostrouchov, Kelly Cho, Yuk-Lam Ho, Edmon Begoli, Junwei Lu, Katherine P. Liao, Tianxi Cai; **Data processing: and Analysis**: Chuan Hong, Everett Rush, Molei Liu, Jiehuan Sun, Aaron Sonabend, Victor M. Castro, Petra Schubert, Vidul A. Panickan, Tianrun Cai, Zeling He, Nicholas Link; **Project administration**: Lauren Costa; **Writing**: Chuan Hong, Everett Rush, Molei Liu, Doudou Zhou, Victor M. Castro, Petra Schubert, Tianxi Cai, Katherine P. Liao; **Guarantors:** Tianxi Cai, Katherine P. Liao; **Approval of final manuscript**: all authors.

## ACKNOWLEDGEMENTS

We would like to acknowledge that this work would not have been possible without the collaboration between VA and the Department of Energy which provided the computing infrastructure necessary to develop and test these approaches at scale using nationwide VA EHR data. We would also like to thank Ms. Hope Cook, SQL Database Administrator, and Mr. Ian Goethert, Data Engineer, for their assistance with optimizing the computing environment required for this study.

This manuscript has been in part co-authored by UT-Battelle, LLC under Contract No. DE-AC05-00OR22725, and under a joint program with the Department of Veterans Affairs under the Million Veteran Project Computational Health Analytics for Medical Precision to Improve Outcomes Now. The United States Government retains and the publisher, by accepting the article for publication, acknowledges that the United States Government retains a nonexclusive, paid-up, irrevocable, world-wide license to publish or reproduce the published form of this manuscript, or allow others to do so, for United States Government purposes. The Department of Energy will provide public access to these results of federally sponsored research in accordance with the DOE Public Access Plan (http://energy.gov/downloads/doe-public-access-plan).

## FUNDING

This research used resources of the Knowledge Discovery Infrastructure at the Oak Ridge National Laboratory, which is supported by the Office of Science of the U.S. Department of Energy under Contract No. DE-AC05-00OR22725.

## Notes

### Competing Interest Statement

The authors have declared no competing interest.

### Author Declarations

The study protocol was approved by the MGB Human Research Committee (IRB00010756). No patient contact occurred in this study which relied on secondary use of data allowing for waiver of informed consent as detailed by 45 CFR 46.116. These activities were approved through the VA Central IRB. They were supported by Million Veteran Program, VA Central IRB 10-02, and approved under VA Central IRB protocol 18-38.

