## Supplemental Files for "Clinical Knowledge Extraction via Sparse Embedding Regression (KESER) with Multi-Center Large Scale Electronic Health Record Data"

This supplementary material provides Implementation details of the feature selection approaches, and additional figures and tables.

**Appendix A. Implementation details of the feature selection approaches**

As we used dropout training in the feature selection procedures including the local regularized regression and the integrative regularized regression, we up-sampled the original training data with 10 folds to reduce the randomness of the results incurred by dropout.

To select the tuning parameter $\lambda$, we split the raw patient-level samples into train and validation sets at each site (11.6:1 split for VA and 1:1 split for MGB), extract SPPMI matrices for the partitioned data set and factorized them into embedding matrices of the same dimensionality. Denote the derived embedding as $\boldsymbol{V}_{train}^{\left( m \right)}$and $\boldsymbol{V}_{valid}^{\left( m \right)}$ for the training and validating set at site $m$, respectively. Then for each target code $w$, we specified a candidate set for the tuning parameter that should be broad enough, and learned the coefficients $B_{wc}^{\left( m \right)}$'s with the training data $\boldsymbol{V}_{train}^{\left( m \right)}$ for each $\lambda\in\Lambda,$ using the local or integrative regularized regression as described in (5) or (6) of our paper. Let $\hat{B}_{wc}^{\left( m \right)}\left( \lambda_{m1},\lambda_{m2} \right)$be the solution corresponding to $\lambda_{m1},\lambda_{m2}$ in the site level feature selection, and $\hat{B}_{wc}^{\left( m \right)}\left( \lambda_{1},\lambda_{2} \right)$be the solution corresponding to $\lambda_{1},\lambda_{2}$ in the integrative regression.

We then chose the parameters minimizing the sum squared loss:

$$\left\| \boldsymbol{V}_{w}^{\left( m \right)}-\sum_{c\in\Omega_{0.05}^{\left( m \right)}} \hat{B}_{wc}^{\left( m \right)}\left( \lambda_{m1},\lambda_{m2} \right)\boldsymbol{V}_{c}^{\left( m \right)} \right\|_{2}^{2}$$

for each $m$ in the local regression and that minimizes

$$\sum_{m=1}^{M} \left\| \boldsymbol{V}_{w}^{\left( m \right)}-\Sigma_{c\in\Omega_{0.05}^{\left( m \right)}}\hat{B}_{wc}^{\left( m \right)}\left( \lambda_{1},\lambda_{2} \right)\boldsymbol{V}_{c}^{\left( m \right)} \right\|_{2}^{2}$$

in the integrative regression. After choosing the optimal parameters, we used it to fit again (5) or (6) with the embedding matrices $\mathbb{V}^{\left( m \right)}$'s derived with the full sample, to obtain the final results.

To speed up our tuning procedure, we separately tune the coefficients for the ridge penalty and sparse penalty. First, we fit ridge regression with penalty coefficients $\lambda_{2}$ or $\lambda_{m2}$ selected by minimizing the sum squared loss on the validation set. Then we take the predicted values output by the tuned ridge regression as a pseudo-outcome to perform sparse or group sparse regression with penalty $\lambda_{2}$ or $\lambda_{m2}$, and again select the parameters minimizing the squared loss on the validation set. To tune each lambda, we initially choose the one among

$${10}^{-5}, {10}^{-5}e^{d}, {10}^{-5}e^{2d}, \ldots, {10}^{-5}e^{1000d},$$

that minimize the sum squared loss on the validation set, where $d=0.01ln10$ so that the candidate set ranges from ${10}^{-5}$ to ${10}^{5}$. Denote the selected parameter as $\lambda_{init}.$ We then choose the parameter among

$$\frac{\lambda_{init}}{4},\frac{\lambda_{init}}{4}+f\lambda_{init},\frac{\lambda_{init}}{4}+2f\lambda_{init},\ldots\frac{\lambda_{init}}{4}+1000f\lambda_{init},$$

where $f=$3/800 so that the candidate set ranges from $\frac{\lambda_{init}}{4}$ to $4\lambda_{init}$.

**Appendix B. Additional figures and tables.**

**Figure S1.** Percentage of variation explained by top d-dimensional eigenvectors at over a range of d at MGB and VA.

(A) MGB

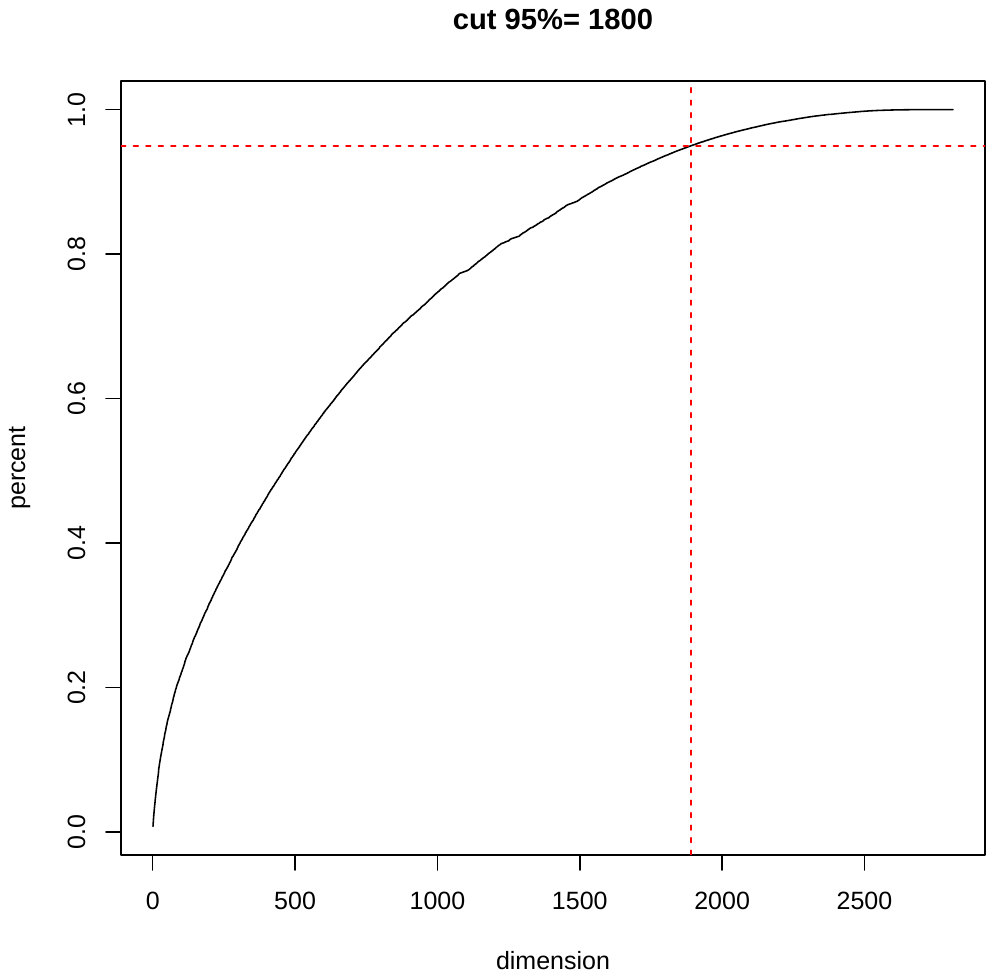

(B) VA.

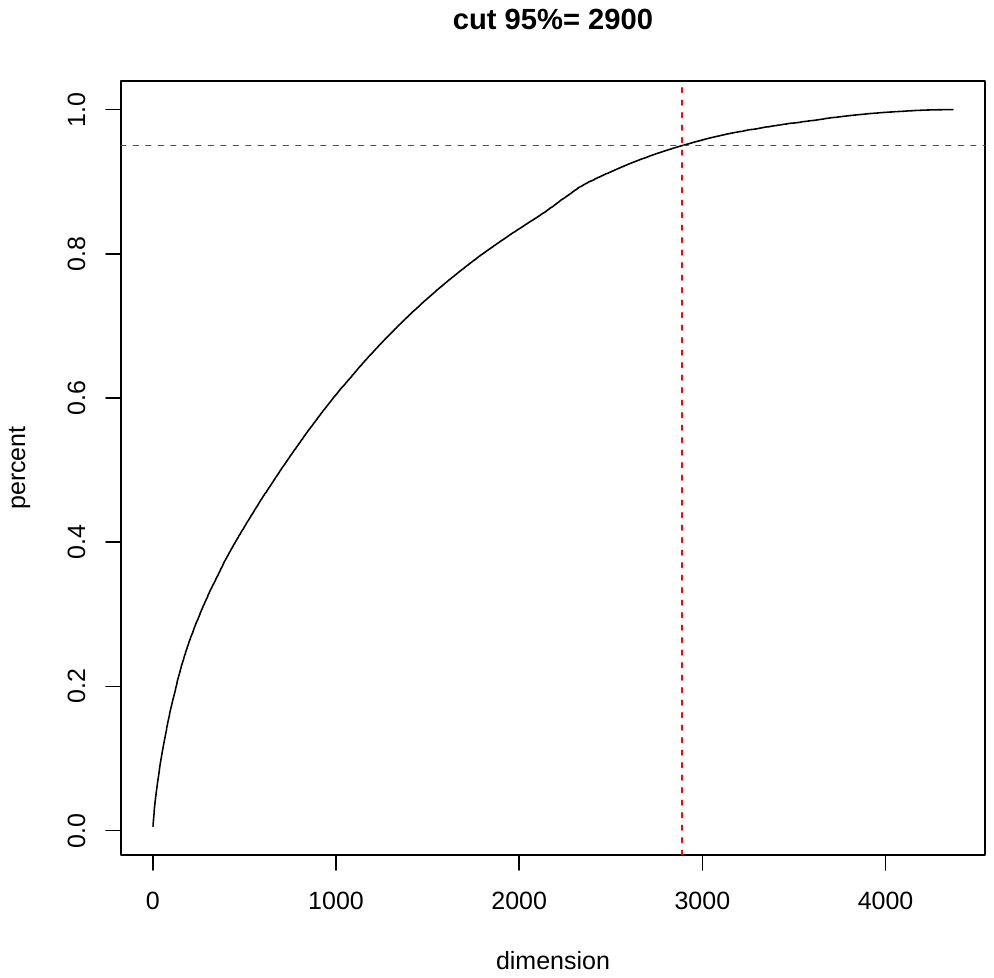

**Figures S2.** KESER_VA_ selected features for Rheumatoid arthritis.

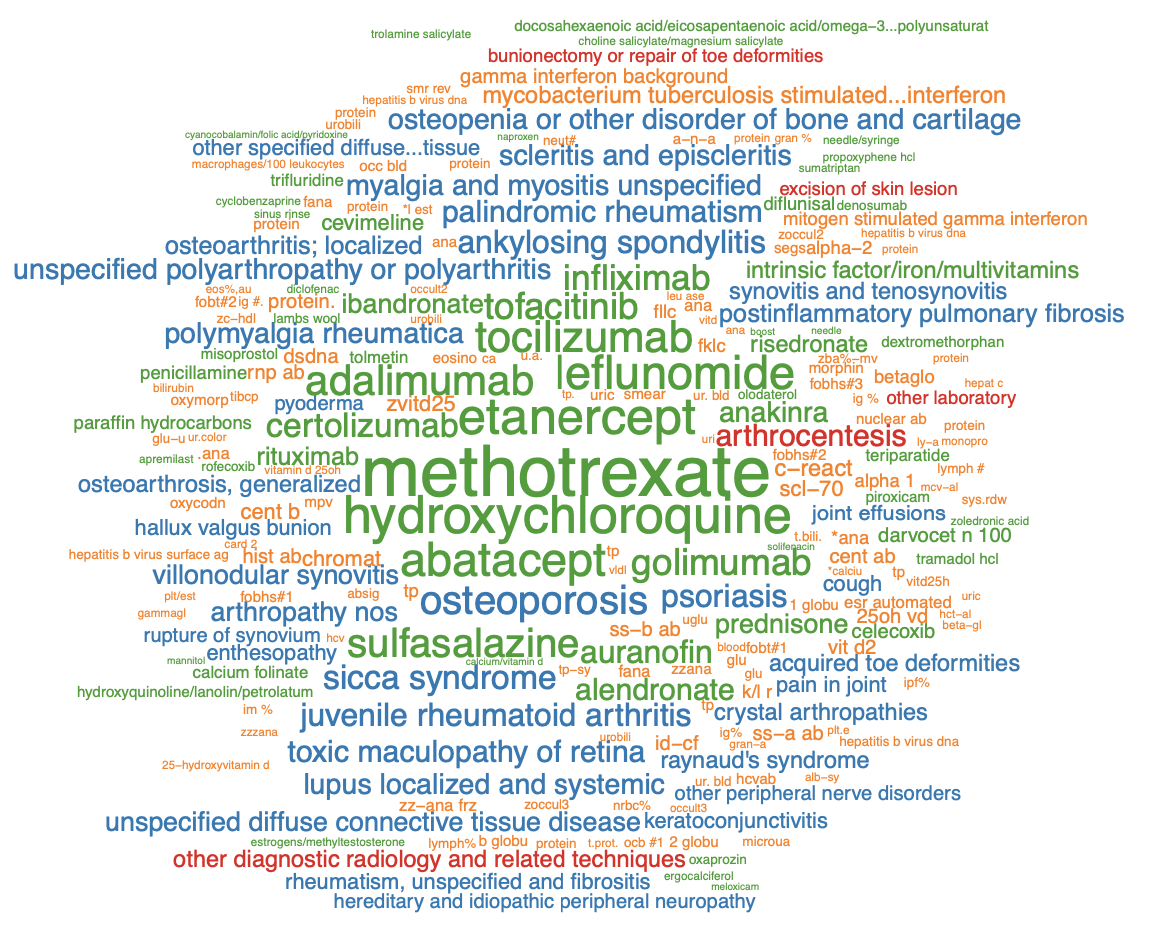

**Figures S3.** KESER selected features for Coronary Artery Disease from MGB and VA.

(A) MGB

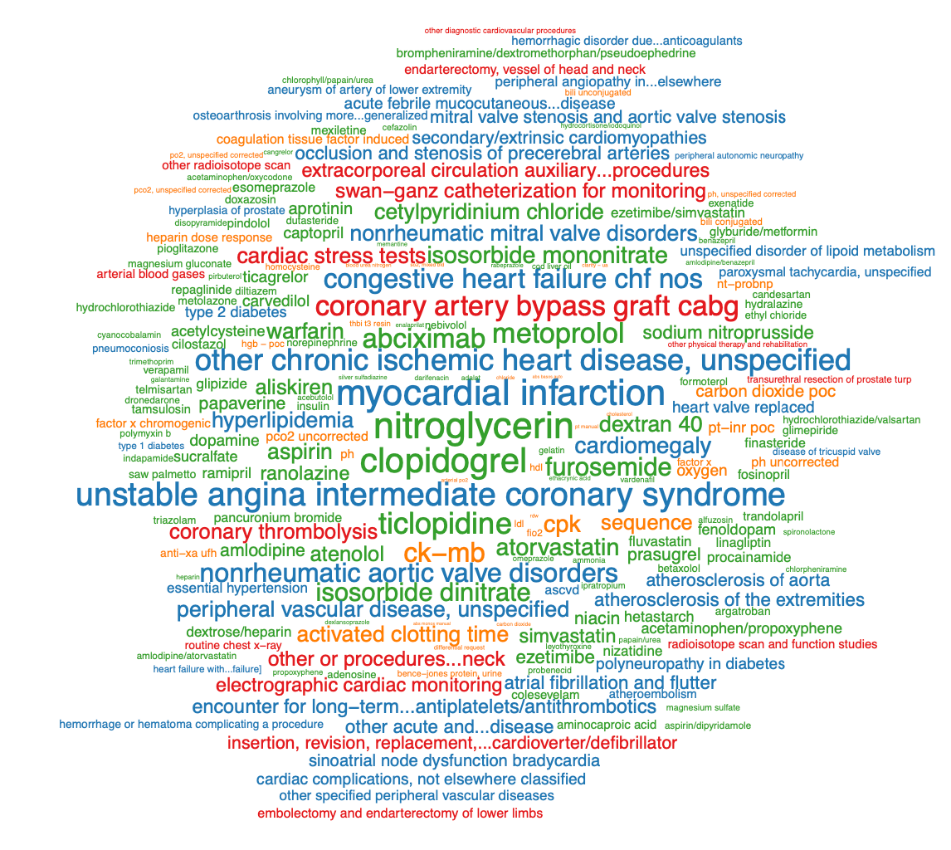

(B) VA

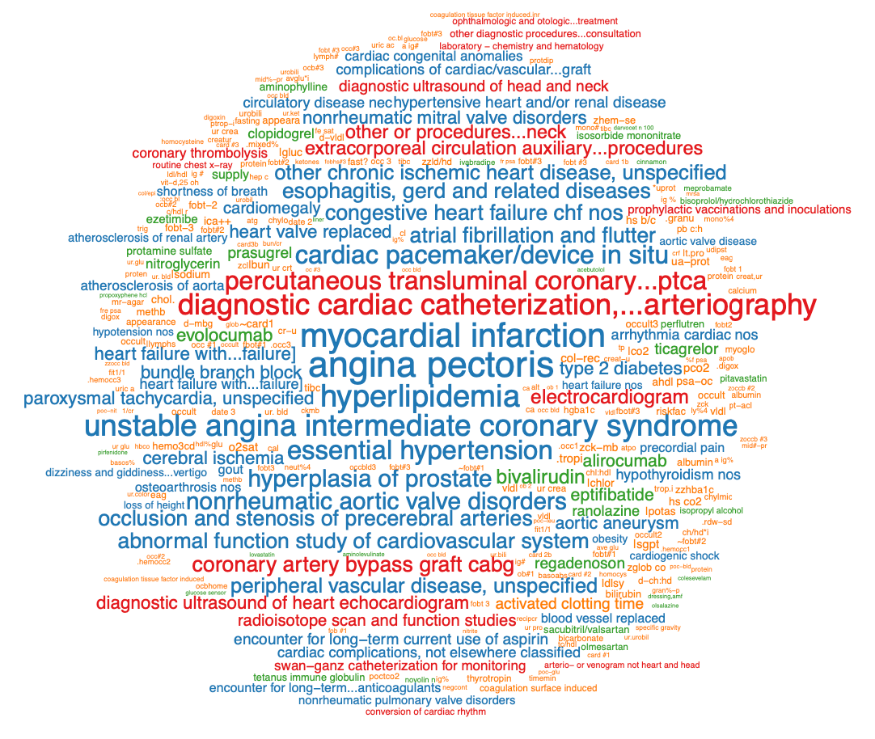

**Figures S4.** KESER selected features for Depression from MGB and VA.

(A) MGB

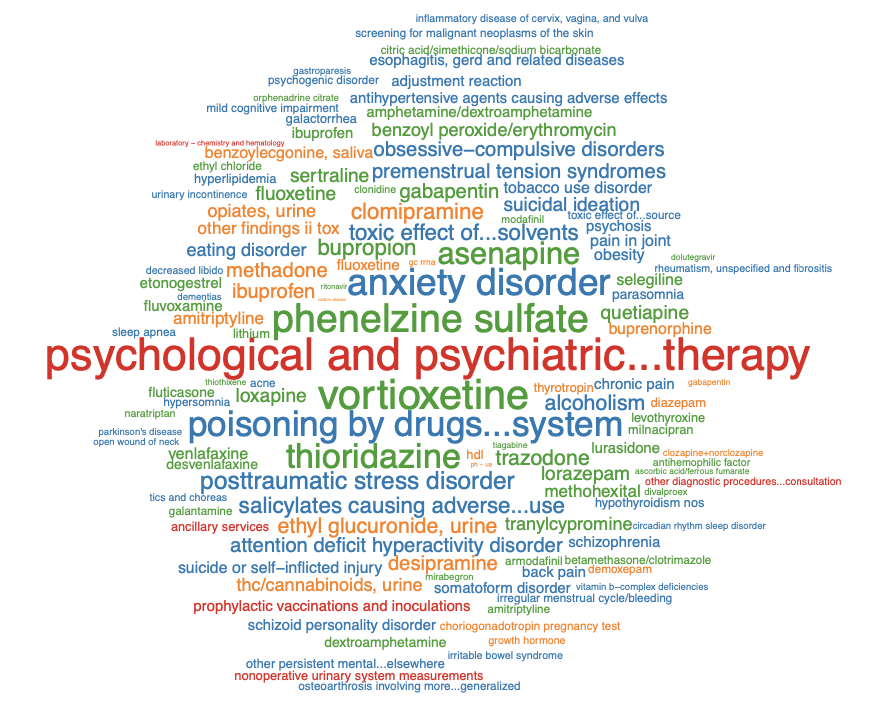

(B) VA

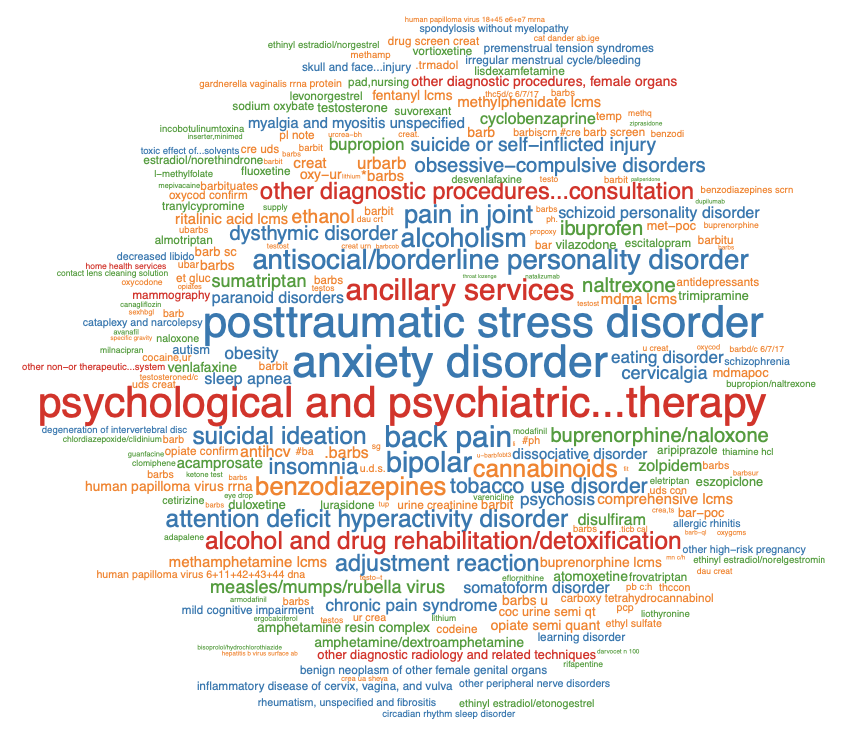

**Figures S5.** KESER selected features for Type 1 diabetes from MGB and VA.

(A) MGB

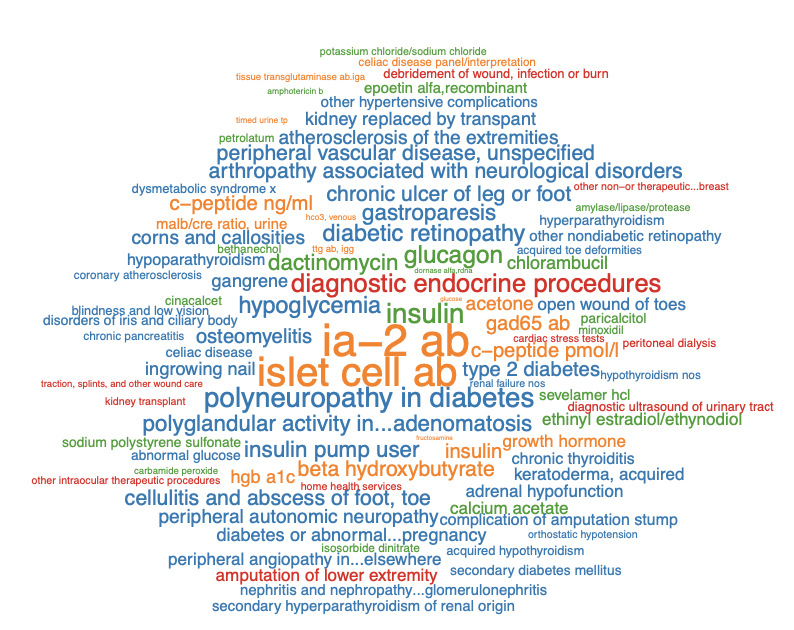

(B) VA

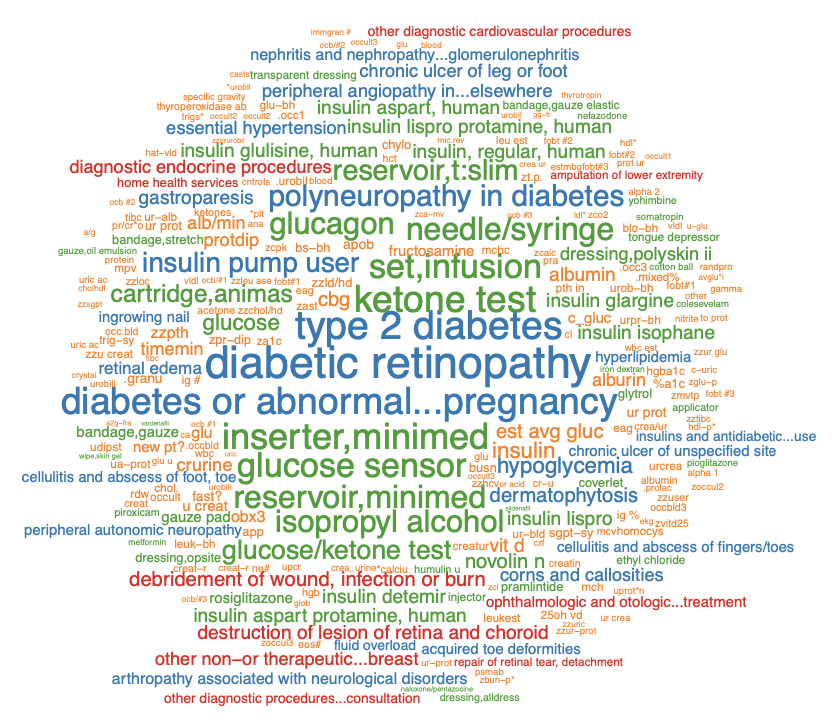

**Figures S6.** KESER selected features for Type 2 diabetes from MGB and VA.

(A) MGB

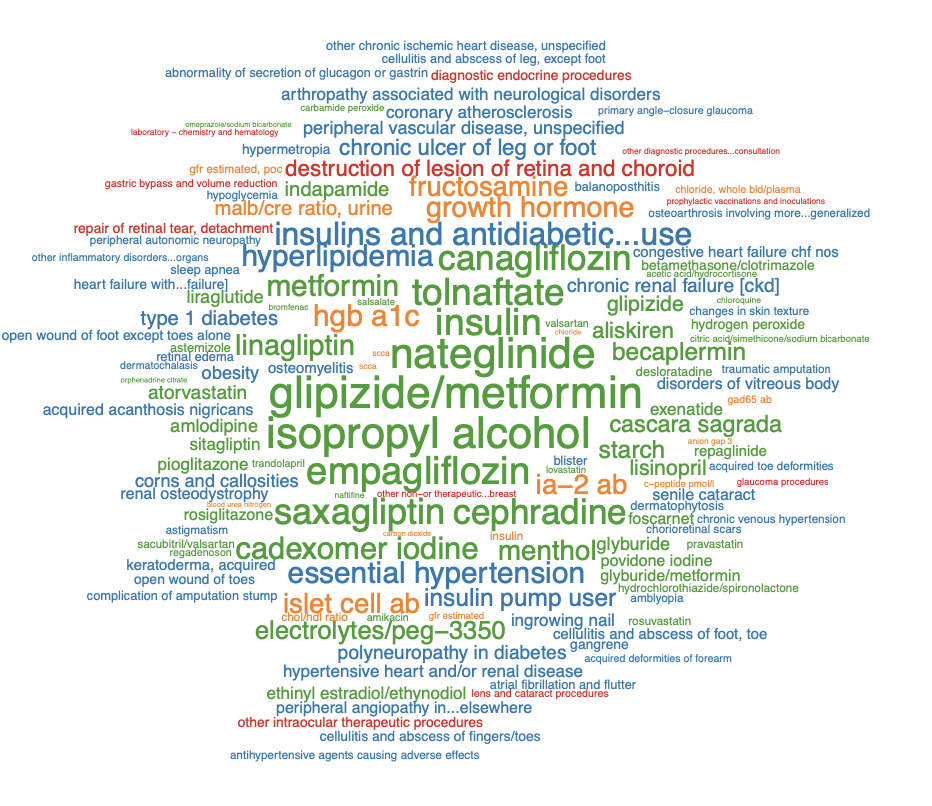

(B) VA

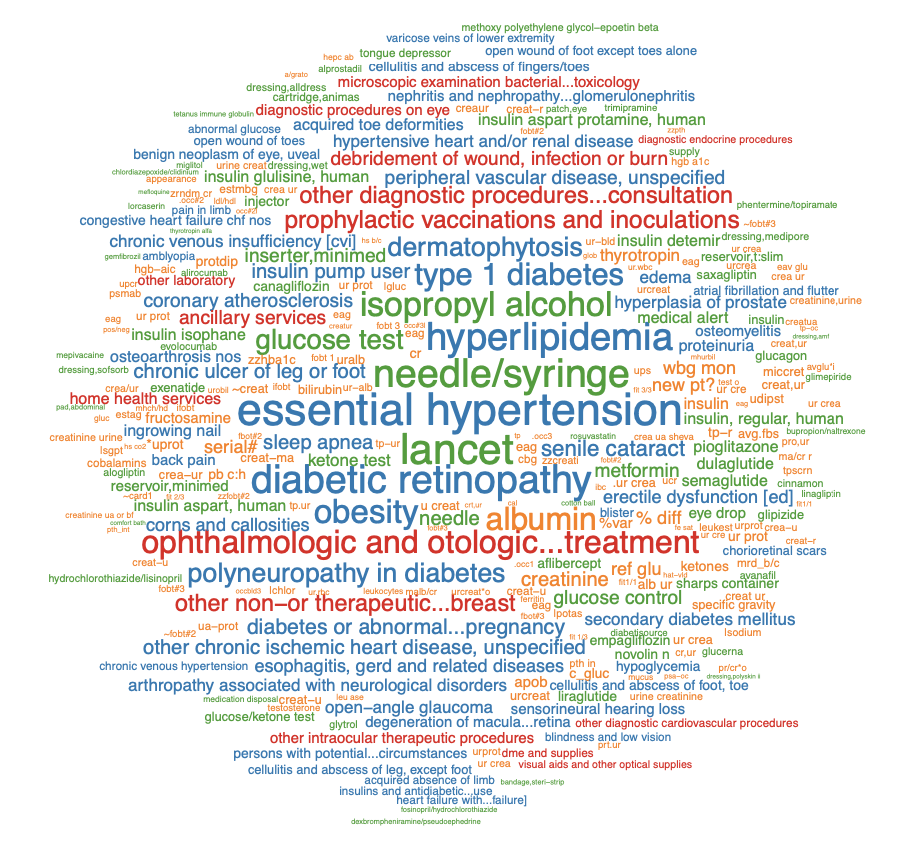

**Figures S7.** KESER selected features for Multiple sclerosis from MGB and VA.

(A) MGB

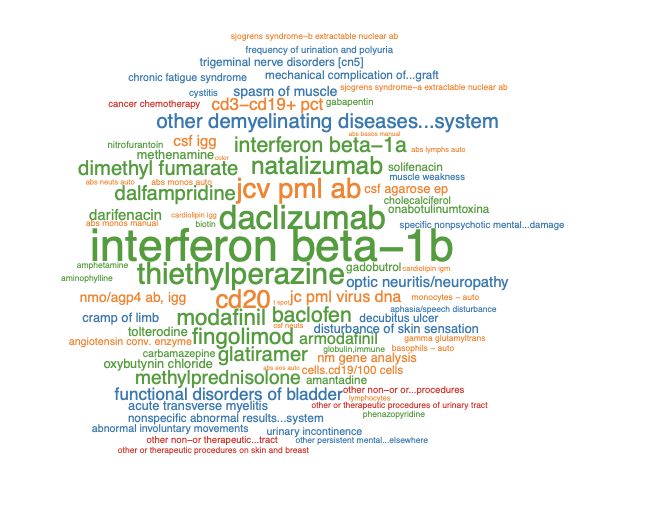

(B) VA

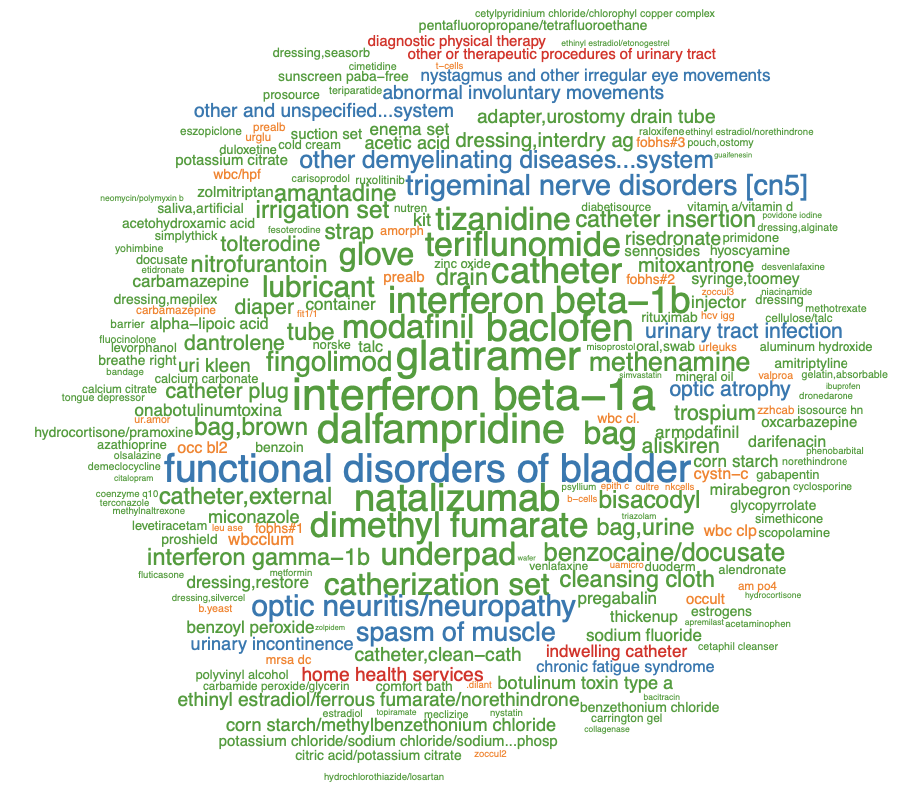

**Figures S8.** KESER selected features for Ulcerative colitis from VA.

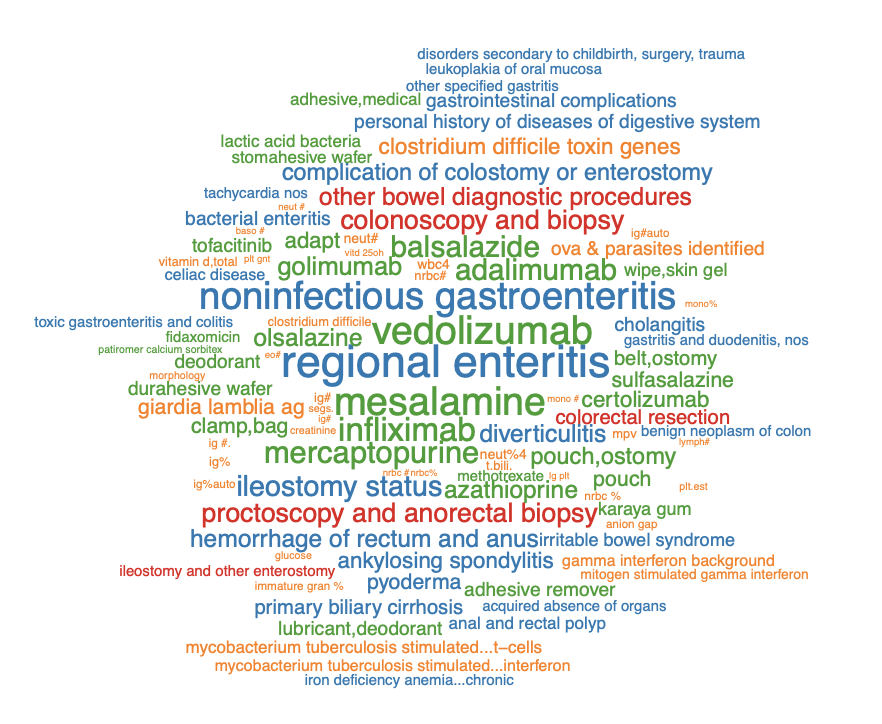

**Figures S9.** KESER selected features for Regional enteritis from MGB and VA.

(A) MGB

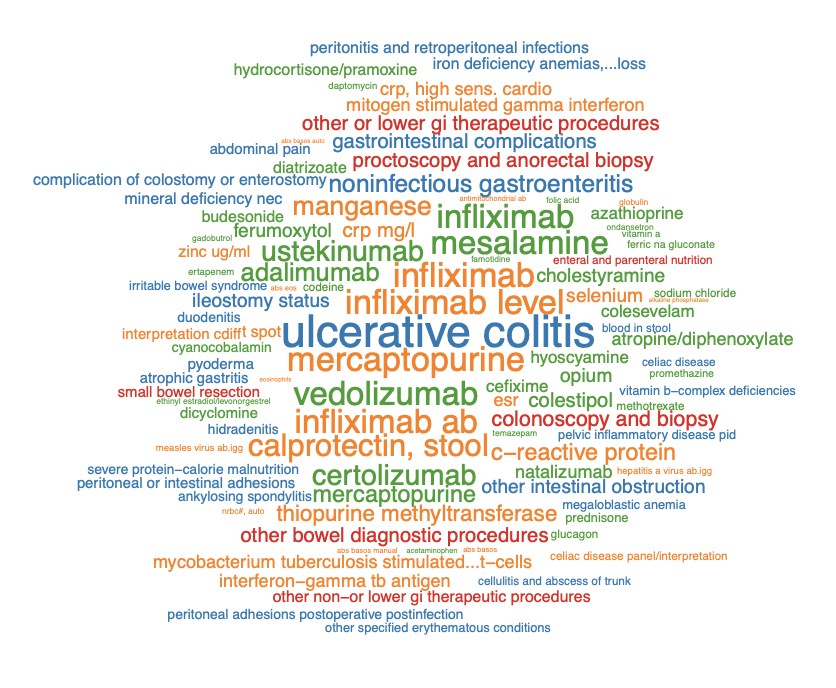

(B) VA

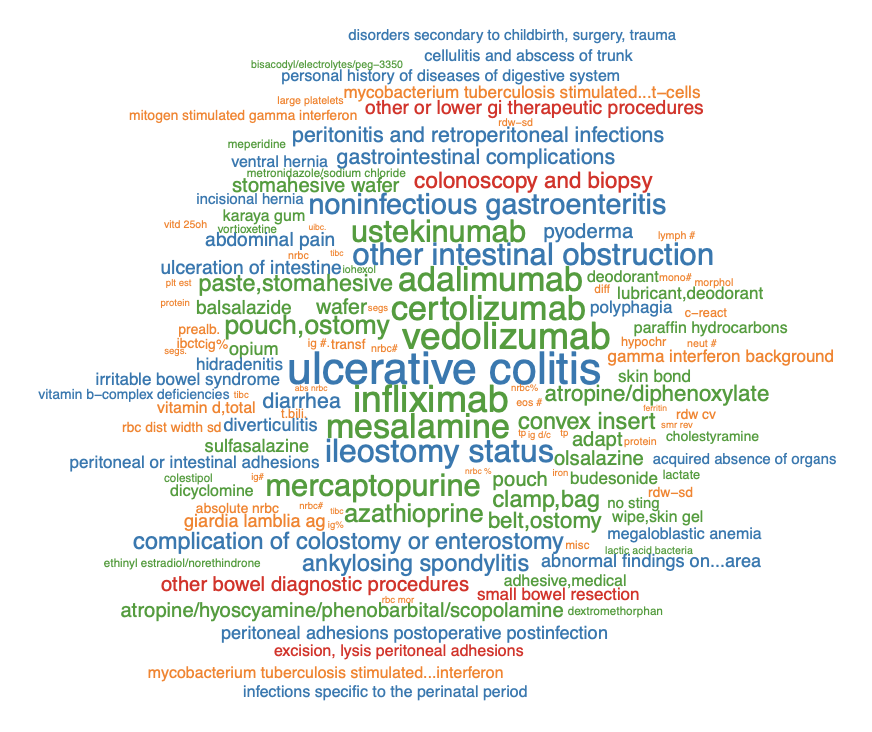

**Figure S10.** Comparison of AUCROCs, AUCPRCs and F-scores with gold standard labels for random forest phenotyping algorithms for 8 diseases using the main PheCode only (PheCode), all features (FULL), SAFE selected features (SAFE), KESER_INT_ selected features using SVD-SPPMI embeddings, KESER_MGB_ selected features using SVD-SPPMI embeddings, as well as KESER_INT_ and KESER_MGB_ selected features based on GloVE embeddings. F-scores are calculated at the cutoff points with the estimated prevalence equal to the population prevalence. The 95% confidence intervals are calculated using bootstrap.

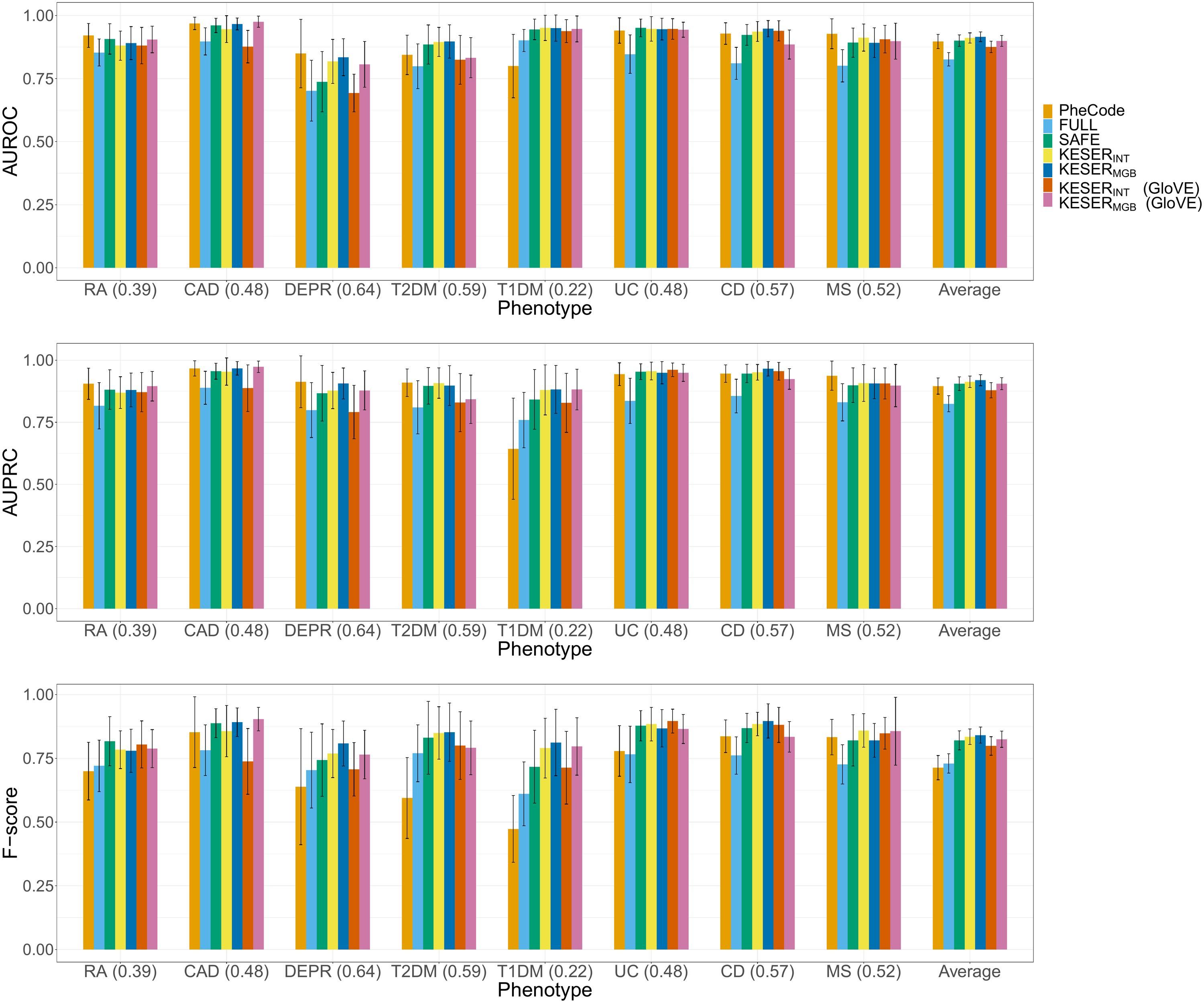

**Figure S11. Features with high cosine similarity with COVID ICD code.**

(a) **MGB**

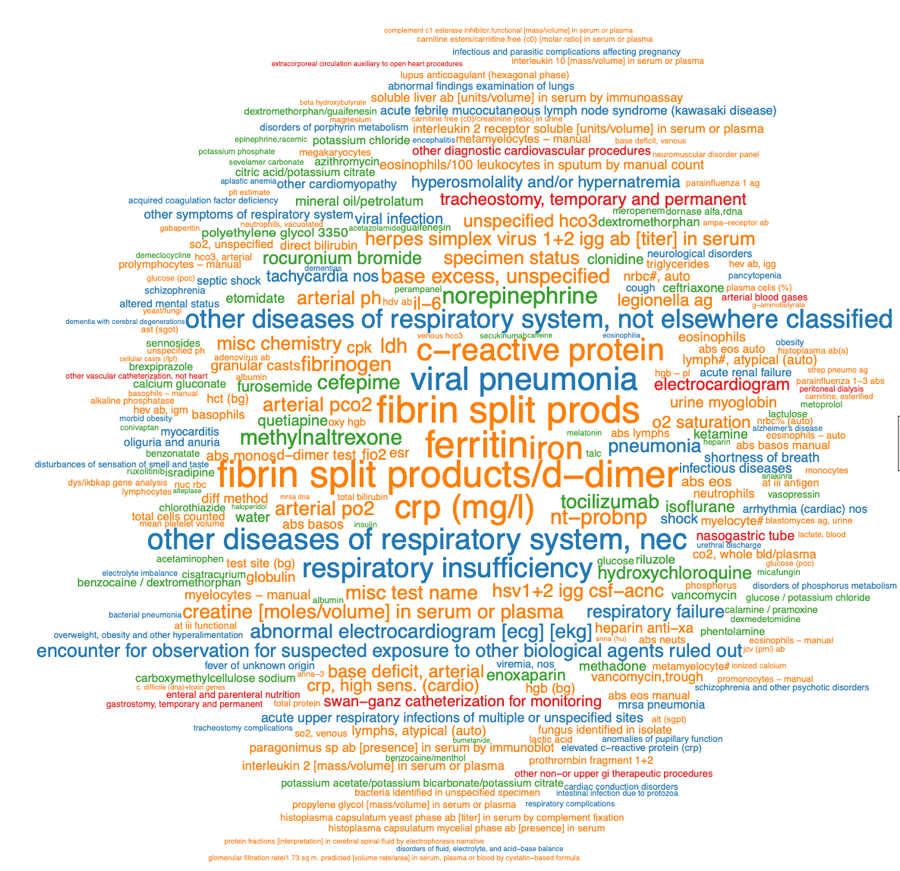

(b) **VA.**

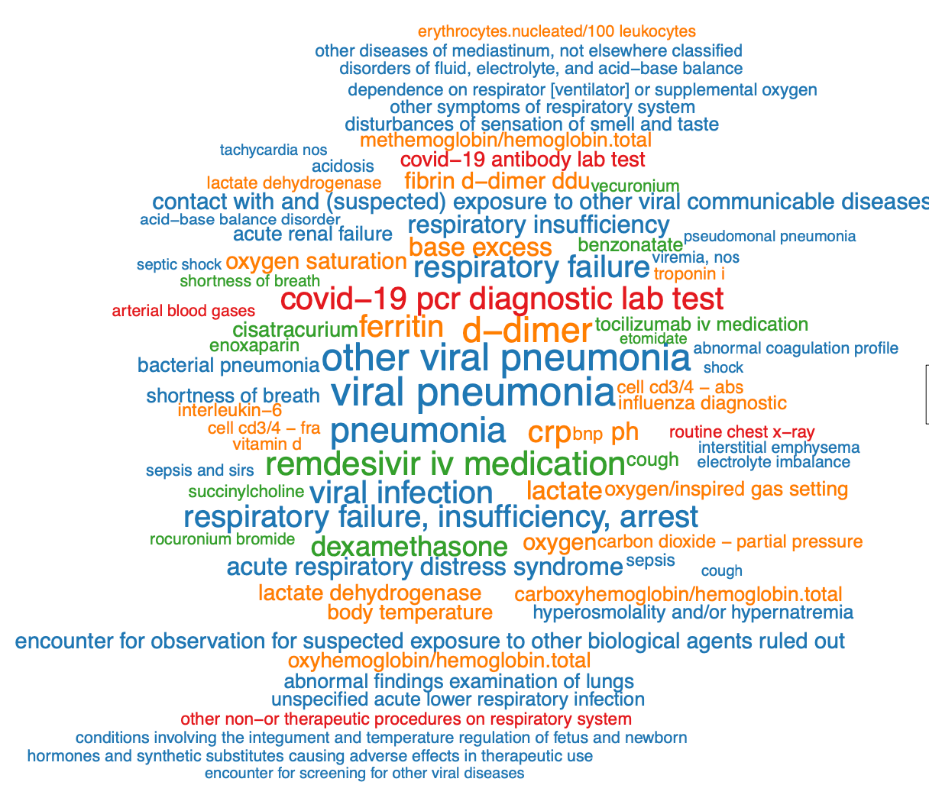

**Table S1.** Known relation pairs from different resources.

| **Usage** | **Entity Pairs** |  | **Relation Type** | **# Pairs** | |
| --- | --- | --- | --- | --- | --- |
|  |  | **Source** |  | **MGB** | **VA** |
| **Optimize dimension for similarity** | **PheCode-PheCode** | **PheCode hierarchy** | **Similar** | **4220** | **4094** |
| **Evaluation** | **PheCode-PheCode** | **Wikipedia** | **Related** | **2306** | **2430** |
|  |  |  | *May Causes* | *243* | *258* |
|  |  |  | *Complications* | *435* | *449* |
|  |  |  | *Symptoms* | *549* | *560* |
|  |  |  | *Risk Factors* | *337* | *342* |
|  |  |  | *Differential Diagnosis* | *439* | *499* |
|  |  |  | *Other* | *90* | *99* |
|  | **PheCode-RxNorm** | **MEDRT, SNOMED-CT, Drug.com** | **Related** | **4416** | **4627** |
|  | **RxNorm-RxNorm** | **SNOMED-CT** | **Similar** | **4002** | **3647** |
|  | **Lab-Lab** | **Manual annotated pairs** | **Similar** | **652** | **426** |

**Table S2.** Sensitivity analysis for choosing dimension *d*, window size *w* and shift parameter *k*.

|  |  |  | **AUC** | | | **TPR (FPR=0.01)** | | | **TPR (FPR=0.05)** | | | **TPR (FPR=0.1)** | | |
| --- | --- | --- | --- | --- | --- | --- | --- | --- | --- | --- | --- | --- | --- | --- |
| **Relation Type** | ***d*** | ***w*** | ***k=1*** | ***k=5*** | ***k=10*** | ***k=1*** | ***k=5*** | ***k=10*** | ***k=1*** | ***k=5*** | ***k=10*** | ***k=1*** | ***k=5*** | ***k=10*** |
| Similar | 100 | 7 | 0.849 | 0.816 | 0.796 | 0.351 | 0.313 | 0.371 | 0.567 | 0.546 | 0.564 | 0.681 | 0.672 | 0.632 |
|  |  | 30 | 0.857 | 0.832 | 0.79 | 0.385 | 0.336 | 0.392 | 0.573 | 0.549 | 0.526 | 0.683 | 0.661 | 0.607 |
|  |  | 60 | 0.861 | 0.823 | 0.794 | 0.37 | 0.351 | 0.366 | 0.583 | 0.567 | 0.528 | 0.694 | 0.654 | 0.59 |
|  | 500 | 7 | 0.845 | 0.817 | 0.801 | 0.423 | 0.452 | 0.4 | 0.628 | 0.651 | 0.612 | 0.71 | 0.705 | 0.675 |
|  |  | 30 | 0.859 | 0.821 | 0.8 | 0.425 | 0.418 | 0.437 | 0.615 | 0.637 | 0.607 | 0.71 | 0.699 | 0.665 |
|  |  | 60 | 0.862 | 0.825 | 0.783 | 0.454 | 0.465 | 0.456 | 0.621 | 0.638 | 0.594 | 0.711 | 0.704 | 0.644 |
|  | *d_95%_* | 7 | 0.828 | 0.788 | 0.764 | 0.394 | 0.361 | 0.359 | 0.586 | 0.616 | 0.602 | 0.683 | 0.675 | 0.653 |
|  |  | 30 | 0.832 | 0.787 | 0.76 | 0.396 | 0.301 | 0.418 | 0.577 | 0.603 | 0.582 | 0.673 | 0.662 | 0.631 |
|  |  | 60 | 0.841 | 0.797 | 0.752 | 0.442 | 0.448 | 0.446 | 0.594 | 0.609 | 0.564 | 0.677 | 0.676 | 0.615 |
| Related | 100 | 7 | 0.794 | 0.739 | 0.7 | 0.1 | 0.094 | 0.131 | 0.357 | 0.322 | 0.294 | 0.511 | 0.454 | 0.399 |
|  |  | 30 | 0.798 | 0.742 | 0.707 | 0.103 | 0.086 | 0.089 | 0.341 | 0.316 | 0.261 | 0.522 | 0.455 | 0.365 |
|  |  | 60 | 0.799 | 0.731 | 0.696 | 0.119 | 0.119 | 0.133 | 0.372 | 0.318 | 0.26 | 0.53 | 0.429 | 0.362 |
|  | 500 | 7 | 0.802 | 0.722 | 0.68 | 0.165 | 0.174 | 0.134 | 0.439 | 0.376 | 0.333 | 0.556 | 0.484 | 0.422 |
|  |  | 30 | 0.804 | 0.711 | 0.68 | 0.14 | 0.15 | 0.134 | 0.408 | 0.363 | 0.329 | 0.557 | 0.459 | 0.421 |
|  |  | 60 | 0.805 | 0.712 | 0.681 | 0.16 | 0.161 | 0.167 | 0.421 | 0.359 | 0.32 | 0.563 | 0.458 | 0.408 |
|  | *d_95%_* | 7 | 0.812 | 0.712 | 0.673 | 0.196 | 0.2 | 0.181 | 0.467 | 0.388 | 0.342 | 0.592 | 0.486 | 0.432 |
|  |  | 30 | 0.814 | 0.71 | 0.678 | 0.158 | 0.177 | 0.148 | 0.446 | 0.383 | 0.322 | 0.583 | 0.46 | 0.418 |
|  |  | 60 | 0.813 | 0.708 | 0.678 | 0.187 | 0.177 | 0.175 | 0.446 | 0.373 | 0.326 | 0.579 | 0.455 | 0.411 |

**Table S3.** Code translation accuracy for VA medication code 🡪 RXNORM and PheCode 🡪 PheCode using embedding dimensions either optimized for AUC (*d_auc_* = 400 at VA, 200 at MGB) or for SNR (*d_snr_* = 1800 at VA, 1000 at MGB).

| **Mapping** | | **Dimensions** | | **Top1** | **Top5** | **Top10** |
| --- | --- | --- | --- | --- | --- | --- |
| **VA** | **MGB** | **VA** | **MGB** |  |  |  |
| VA medication code | RxNorm | 400 | 200 | 0.382 | 0.665 | 0.777 |
|  |  | 1800 | 1000 | 0.394 | 0.669 | 0.793 |
| PheCode | PheCode | 400 | 200 | 0.421 | 0.735 | 0.844 |
|  |  | 1800 | 1000 | 0.385 | 0.717 | 0.806 |

**Table S4.** AUC and sensitivity at FPR = 0.01, 0.05 and 0.1 of cosine similarity in detecting (a) known similar pairs (RxNorm-RxNorm and Lab-Lab); and (b) related pairs across different types of relationships.

(a) Similar pairs

| **Entity Pair** | **Method** | **Dimension** | **AUC** | | **Sensitivity** | | | | | |
| --- | --- | --- | --- | --- | --- | --- | --- | --- | --- | --- |
|  |  |  |  |  | **FPR = 0.01** | | **FPR = 0.05** | | **FPR = 0.1** | |
|  |  |  | **MGB** | **VA** | **MGB** | **VA** | **MGB** | **VA** | **MGB** | **VA** |
| RxNorm-RxNorm | GloVe | 50 | 0.790 | 0.757 | 0.180 | 0.146 | 0.363 | 0.344 | 0.470 | 0.477 |
|  |  | 100 | 0.797 | 0.763 | 0.177 | 0.142 | 0.350 | 0.341 | 0.460 | 0.460 |
|  | PPMI | 100 | 0.749 | 0.793 | 0.110 | 0.191 | 0.326 | 0.328 | 0.439 | 0.474 |
|  |  | 500 | 0.775 | 0.841 | 0.210 | 0.298 | 0.430 | 0.520 | 0.541 | 0.638 |
|  |  | *d*_s_*_nr_* (1000,1800) | 0.775 | 0.830 | 0.223 | 0.247 | 0.430 | 0.457 | 0.533 | 0.602 |
|  |  | *d_auc_*(300,500) | 0.773 | 0.841 | 0.174 | 0.298 | 0.396 | 0.520 | 0.533 | 0.638 |
|  |  | *d_95%_*(1800,2900) | 0.772 | 0.825 | 0.202 | 0.242 | 0.424 | 0.449 | 0.541 | 0.585 |
| Lab-Lab | GloVe | 50 | 0.949 | 0.962 | 0.670 | 0.627 | 0.844 | 0.897 | 0.902 | 0.932 |
|  |  | 100 | 0.955 | 0.947 | 0.689 | 0.641 | 0.877 | 0.836 | 0.903 | 0.899 |
|  | PPMI | 100 | 0.913 | 0.922 | 0.670 | 0.345 | 0.791 | 0.669 | 0.813 | 0.817 |
|  |  | 500 | 0.910 | 0.934 | 0.724 | 0.507 | 0.785 | 0.838 | 0.804 | 0.906 |
|  |  | *d*_s_*_nr_* (1000,1800) | 0.900 | 0.910 | 0.724 | 0.568 | 0.775 | 0.805 | 0.807 | 0.873 |
|  |  | *d_auc_*(300,500) | 0.905 | 0.934 | 0.736 | 0.507 | 0.790 | 0.838 | 0.805 | 0.906 |
|  |  | *d_95%_*(1800,2900) | 0.901 | 0.910 | 0.729 | 0.531 | 0.778 | 0.826 | 0.813 | 0.883 |
| (b) Related pairs | |  | |  | |  | |  | | |
| **Entity Pair** | **Method** | **Dimension** | **AUC** | | **Sensitivity** | | | | | |
|  |  |  |  |  | **FPR = 0.01** | | **FPR = 0.05** | | **FPR = 0.1** | |
|  |  |  | **MGB** | **VA** | **MGB** | **VA** | **MGB** | **VA** | **MGB** | **VA** |
| PheCode-PheCode | GloVe | 50 | 0.947 | 0.838 | 0.439 | 0.290 | 0.745 | 0.464 | 0.867 | 0.594 |
| *May cause* |  | 100 | 0.948 | 0.870 | 0.396 | 0.307 | 0.773 | 0.522 | 0.871 | 0.621 |
|  | PPMI | 100 | 0.901 | 0.868 | 0.424 | 0.326 | 0.654 | 0.550 | 0.737 | 0.628 |
|  |  | 500 | 0.913 | 0.913 | 0.519 | 0.500 | 0.658 | 0.702 | 0.753 | 0.771 |
|  |  | *d*_s_*_nr_* (1800,2800) | 0.924 | 0.925 | 0.502 | 0.519 | 0.774 | 0.694 | 0.819 | 0.810 |
|  |  | *d_auc_*(1800,2300) | 0.924 | 0.925 | 0.502 | 0.516 | 0.774 | 0.698 | 0.819 | 0.810 |
|  |  | *d_95%_*(1800,2900) | 0.924 | 0.925 | 0.502 | 0.519 | 0.774 | 0.694 | 0.819 | 0.810 |
| PheCode-PheCode | GloVe | 50 | 0.875 | 0.799 | 0.331 | 0.245 | 0.595 | 0.361 | 0.671 | 0.470 |
| *complications* |  | 100 | 0.881 | 0.825 | 0.362 | 0.263 | 0.554 | 0.388 | 0.709 | 0.538 |
|  | PPMI | 100 | 0.859 | 0.815 | 0.241 | 0.241 | 0.540 | 0.419 | 0.648 | 0.526 |
|  |  | 500 | 0.880 | 0.870 | 0.257 | 0.303 | 0.628 | 0.566 | 0.726 | 0.655 |
|  |  | *d*_s_*_nr_* (1800,2800) | 0.884 | 0.865 | 0.248 | 0.285 | 0.653 | 0.521 | 0.743 | 0.637 |
|  |  | *d_auc_*(1800,2300) | 0.884 | 0.865 | 0.248 | 0.285 | 0.653 | 0.532 | 0.743 | 0.637 |
|  |  | *d_95%_*(1800,2900) | 0.884 | 0.866 | 0.248 | 0.287 | 0.653 | 0.521 | 0.743 | 0.637 |
| PheCode-PheCode | GloVe | 50 | 0.894 | 0.814 | 0.234 | 0.217 | 0.534 | 0.460 | 0.723 | 0.519 |
| *differential* |  | 100 | 0.889 | 0.854 | 0.175 | 0.278 | 0.558 | 0.513 | 0.727 | 0.593 |
| *diagnosis* | PPMI | 100 | 0.901 | 0.887 | 0.324 | 0.364 | 0.585 | 0.538 | 0.741 | 0.661 |
|  |  | 500 | 0.896 | 0.912 | 0.388 | 0.461 | 0.678 | 0.632 | 0.754 | 0.770 |
|  |  | *d*_s_*_nr_* (1800,2800) | 0.898 | 0.910 | 0.419 | 0.482 | 0.670 | 0.714 | 0.750 | 0.766 |
|  |  | *d_auc_*(1800,2300) | 0.898 | 0.909 | 0.419 | 0.471 | 0.670 | 0.696 | 0.750 | 0.773 |
|  |  | *d_95%_*(1800,2900) | 0.898 | 0.909 | 0.419 | 0.482 | 0.670 | 0.714 | 0.750 | 0.768 |
| PheCode-PheCode | GloVe | 50 | 0.834 | 0.744 | 0.175 | 0.147 | 0.421 | 0.349 | 0.598 | 0.451 |
| *risk factors* |  | 100 | 0.836 | 0.775 | 0.194 | 0.144 | 0.448 | 0.373 | 0.607 | 0.469 |
|  | PPMI | 100 | 0.760 | 0.687 | 0.092 | 0.096 | 0.329 | 0.211 | 0.445 | 0.272 |
|  |  | 500 | 0.806 | 0.742 | 0.148 | 0.137 | 0.457 | 0.278 | 0.531 | 0.395 |
|  |  | *d*_s_*_nr_* (1800,2800) | 0.816 | 0.760 | 0.160 | 0.184 | 0.463 | 0.336 | 0.611 | 0.442 |
|  |  | *d_auc_*(1800,2300) | 0.816 | 0.762 | 0.160 | 0.181 | 0.463 | 0.342 | 0.611 | 0.442 |
|  |  | *d_95%_*(1800,2900) | 0.816 | 0.759 | 0.160 | 0.184 | 0.463 | 0.336 | 0.611 | 0.442 |
| PheCode-PheCode | GloVe | 50 | 0.870 | 0.804 | 0.278 | 0.210 | 0.520 | 0.361 | 0.701 | 0.513 |
| *symptoms* |  | 100 | 0.869 | 0.834 | 0.264 | 0.203 | 0.540 | 0.431 | 0.703 | 0.574 |
|  | PPMI | 100 | 0.860 | 0.849 | 0.134 | 0.198 | 0.358 | 0.421 | 0.649 | 0.567 |
|  |  | 500 | 0.874 | 0.877 | 0.139 | 0.204 | 0.412 | 0.545 | 0.706 | 0.669 |
|  |  | *d*_s_*_nr_* (1800,2800) | 0.881 | 0.878 | 0.212 | 0.234 | 0.531 | 0.559 | 0.745 | 0.697 |
|  |  | *d_auc_*(1800,2300) | 0.881 | 0.877 | 0.212 | 0.228 | 0.531 | 0.573 | 0.745 | 0.699 |
|  |  | *d_95%_*(1800,2900) | 0.881 | 0.878 | 0.212 | 0.234 | 0.531 | 0.561 | 0.745 | 0.699 |
| PheCode-RxNorm | GloVe | 50 | 0.859 | 0.804 | 0.240 | 0.158 | 0.496 | 0.374 | 0.608 | 0.506 |
|  |  | 100 | 0.863 | 0.817 | 0.254 | 0.217 | 0.493 | 0.421 | 0.625 | 0.533 |
|  | PPMI | 100 | 0.840 | 0.802 | 0.179 | 0.198 | 0.499 | 0.398 | 0.622 | 0.528 |
|  |  | 500 | 0.848 | 0.839 | 0.352 | 0.330 | 0.584 | 0.557 | 0.678 | 0.652 |
|  |  | *d*_s_*_nr_* (1800,2800) | 0.850 | 0.853 | 0.435 | 0.379 | 0.608 | 0.600 | 0.685 | 0.697 |
|  |  | *d_auc_*(1800,2300) | 0.850 | 0.853 | 0.435 | 0.378 | 0.608 | 0.600 | 0.685 | 0.696 |
|  |  | *d_95%_*(1800,2900) | 0.850 | 0.853 | 0.435 | 0.378 | 0.608 | 0.601 | 0.685 | 0.696 |
